# Genomic epidemiology of *S. aureus* isolated from bloodstream infections in South America during 2019 supports regional surveillance

**DOI:** 10.1101/2022.12.20.22283725

**Authors:** Sabrina Di Gregorio, Jesús Vielma, María Sol Haim, Lucía Rago, Josefina Campos, Mihir Kekre, Monica Abrudan, Àngela Famiglietti, Liliana Fernandez Canigia, Gabriela Rubinstein, Martha Helena von Specht, Melina Herrera, Carolina Aro, Marcelo Galas, Norah Balderrama Yarhui, Agnes Figueiredo, Nilton Lincopan, Miryan Falcon, Rosa Guillén, Teresa Camou, Gustavo Varela, David M. Aanensen, Silvia Argimón, Marta Mollerach, StaphNET-SA consortium

## Abstract

*Staphylococcus aureus* remains one of the leading causes of infections worldwide and a common cause of bacteremia. However, studies documenting the epidemiology of *S. aureus* in South America (SA) using genomics are scarce. We hereby report on the largest to date genomic epidemiology study of both methicillin-resistant *S. aureus* (MRSA) and methicillin-susceptible *S. aureus* (MSSA) in SA, conducted by the StaphNET-SA network. We characterised 404 genomes recovered from a prospective observational study of *S. aureus* bacteremia in 58 hospitals from Argentina, Bolivia, Brazil, Paraguay and Uruguay between April and October 2019.

We show that a minority of *S. aureus* isolates are phenotypically multi-drug resistant (5.2%), but more than a quarter are resistant to macrolide-lincosamide-streptogramin B (MLSb). MSSA were more genetically diverse than MRSA. Lower rates of associated antimicrobial resistance in CA-MRSA vs HA-MRSA were found in association with three *S. aureus* genotypes dominating the MRSA population: CC30-MRSA-IVc-*t019*-*lukS/F-PV+*, CC5-MRSA-IV-*t002*-*lukS/F-PV-*, and CC8-MRSA-IVc-*t008*-*lukS/F-PV+*-COMER+. These are historically from a CA origin, carry on average less AMR determinants, and often lack key virulence genes.

Surprisingly, CC398-MSSA-*t1451*-*lukS/F-PV-* related to the CC398 human-associated lineage is widely disseminated throughout the region, and is described here for the first time as the most prevalent MSSA lineage in SA. Moreover, CC398 strains carrying *ermT* and *sh_fabI* (related to triclosan resistance) were recovered from both CA and HA origin, and are largely responsible for the MLSb rates of MSSA strains (inducible iMLSb phenotype).

The frequency of MRSA and MSSA lineages differed between countries but the most prevalent *S. aureus* genotypes are high-risk clones widespread in the South American region without clear country-specific phylogeographic structure. Therefore our findings underscore the need for continuous genomic surveillance by regional networks such as StaphNET-SA.

**Impact statement:** *S. aureus* is a common cause of bacteremia, a serious life threatening disease, and the second leading pathogen for deaths associated with resistance in 2019. However, genomic surveillance of *S. aureus* causing invasive infections in South America is limited. Previous surveillance studies have focused on the dissemination of MRSA with increasing AMR and/or virulence, but have not characterised MSSA in detail.

Here, we show the results of a prospective observational study of genomic surveillance of *S. aureus* causing bacteremia conducted in South America during 2019 by the StaphNET-SA network.

Our study reveals that in 2019 most bloodstream infections were caused by successful MRSA lineages of community origin, generally not MDR, and lacking key virulence genes in some cases. Importantly, we also describe here for the first time CC398-MSSA-*t1451* as the most prevalent and widely disseminated MSSA clone causing bacteraemia in the region during 2019. This human adapted clone, present both in the community and hospital environment, carries a gene conferring resistance against an antiseptic widely used in our region, and is largely responsible for the increasing resistance rates to erythromycin and clindamycin observed in MSSA.

We also show evidence of readily transmission of the most prevalent MRSA and MSSA high-risk clones across country borders, which highlights the need for continuous genomic surveillance by regional networks such as StaphNET-SA.

**Data Summary:** All supporting data, code and protocols have been provided within the article or through supplementary data files. Five supplementary figures and five supplementary tables are available with the online version of this article.

Sequence read files for all samples used in this study have been deposited in the European Nucleotide Archive under the project accession number PRJEB37318. Individual accession numbers for each sample are also detailed in microreact_project: https://microreact.org/project/staphnet-sa-1st-survey. Genome assemblies are available via Pathogenwatch https://pathogen.watch/collection/jz7rcy1zv0sk-staphnet-sa-first-survey.

## Introduction

*Staphylococcus aureus* causes multiple types of pathologies ranging from moderately severe skin infections, food poisoning, to fatal pneumonia and sepsis. Worldwide, it is one of the most frequently isolated microorganisms in both nosocomial and community-acquired infections associated with high morbidity and mortality [1] and the second leading pathogen for deaths associated with resistance in 2019 [2].

The emergence and dissemination of methicillin resistant *S. aureus* (MRSA) and with additional resistance to other antimicrobial agents is a serious problem both in the hospital (HA-MRSA) and the community (CA-MRSA) environment [3]. Nevertheless, the distinction between community and hospital strains has been blurred and CA-MRSA and HA-MRSA strains have been described as causing outbreaks in both settings [4, 5].

The success of *S. aureus* as human pathogen relies not only on its ability to develop resistance to most antimicrobial agents introduced into the clinical practice, but also on its capacity to produce a diverse set of virulence determinants, including components that enable host colonisation and a variety of toxins and immune evasion factors. Different epidemic *S. aureus* lineages represent a serious threat to public health around the world. These high-risk clones (HRCs) may combine greater virulence or transmission potential with resistance to multiple antimicrobial families [6], and their prevalence usually changes over time and geography.

This scenario is also true in South America, where the prevalence of MRSA was reported to be around 50% in most countries [7]. A regional study performed between 2011-2014, revealed that the main MRSA clones causing bacteremia belonged to the CC5, CC8 and CC30 clonal complexes [8]. ST5-MRSA-I (Cordobes/Chilean clone) and ST105-MRSA-II replaced ST239-MRSA-III (Brazilian clone) as the prevalent HA-MRSA epidemic clones. Meanwhile, ST30-MRSA-IV and ST5-MRSA-IV were the main genotypes associated with CA-MRSA in the southern cone of the region [8, 9]. In contrast, ST8-MRSA-IV, related to the hypervirulent clone USA300, also known as USA300 Latin American variant or South American Epidemic (USA300-LV or USA300-SAE), was found predominant in Colombia, Ecuador and Venezuela [8, 10].

Traditionally, surveillance programmes focus on MRSA, but efforts are needed in order to understand the epidemiology of MSSA infections, which are also of high burden, increasing in prevalence [11–13], and can give rise to MRSA. Clonal replacement appears to be a common phenomenon, and continuous surveillance is crucial to identify changes in the molecular epidemiology of both MRSA and MSSA. Genomic epidemiology using whole-genome sequencing (WGS) is a powerful tool for surveillance programmes and can provide valuable information on the emergence of HRCs, antibiotic resistance mechanisms and virulence determinants [14]. Still, studies documenting the epidemiology of *S. aureus* in Latin America using genomics are scarce [8–10, 15].

We established a network for collaborative surveillance that can characterise the geographic and temporal dynamics of MSSA and MRSA and their epidemic patterns in South American countries (StaphNET-SA network). The aim of this study was to describe the circulating *S. aureus* clones in the region during 2019 using WGS, to compare them with the global population, and to develop regional capabilities for the sequencing and bioinformatic analysis of whole genomes of *S. aureus* in South America.

## Material and Methods

### Ethics statement

The present study was approved by the Ethics Committee of the School of Pharmacy and Biochemistry of the University of Buenos Aires (RES (CD) N°818-19 CEIC - FFYB-UBA), and by the Teaching and Research and / or Ethics Committees from the participating Institutions that have this requirement.

The study was non-interventional, and cases were managed clinically both in terms of diagnosis and treatment according to standard care guidelines. Patients’ personal identifiable information (name, dob, address) was not provided for this study. Informed written consent was not required, since the information was anonymized and the medical management and the diagnostic procedures were not affected.

### Bacterial isolates and study design

A prospective observational study of genomic surveillance of *S. aureus* bacteremia was conducted in 58 participating hospitals from Argentina, Bolivia, Brazil, Paraguay, and Uruguay during 2019.

Each participating centre aimed to collect the first five successive MSSA and the first five successive MRSA primary isolates (always from different individuals) obtained from blood cultures between April and October 2019. When a hospital was unable to collect five MRSA or MSSA isolates within the sampling period, the quota of ten could be reached by submitting additional MSSA or MRSA isolates, respectively **(Table 1, Suppl Fig. 1)**. This study protocol was previously employed for two European structured surveys of *S. aureus* [16, 17].

**Table 1.** Summary of collected isolates by country. CA=Community Acquired. HA=Hospital Acquired. ND=Data not available

| Country | Collected strains | Hospitals (n) | MRSA |  |  | Total MRSA | MSSA |  |  | Total MSSA | Total |
| --- | --- | --- | --- | --- | --- | --- | --- | --- | --- | --- | --- |
|  |  |  | CA | HA | ND |  | CA | HA | ND |  |  |
| Argentina | 10 (5 MRSA / 5 MSSA) | 9 | 25 | 20 | 0 | 45 | 17 | 28 | 0 | 45 | 90 |
|  | 10 (≠ 5/5) | 7 | 11 | 15 | 1 | 27 | 17 | 26 | 0 | 43 | 70 |
|  | <10 | 11 | 14 | 6 | 0 | 20 | 23 | 25 | 1 | 49 | 69 |
|  | Total | 27 | 50 | 41 | 1 | 92 | 57 | 79 | 1 | 137 | 229 |
| Bolivia | 10 (5 MRSA / 5 MSSA) | 4 | 9 | 11 | 0 | 20 | 14 | 6 | 0 | 20 | 40 |
|  | 10 (≠ 5/5) | 1 | 3 | 0 | 0 | 3 | 5 | 2 | 0 | 7 | 10 |
|  | <10 | 4 | 6 | 2 | 0 | 8 | 5 | 2 | 0 | 7 | 15 |
|  | Total | 9 | 18 | 13 | 0 | 31 | 24 | 10 | 0 | 34 | 65 |
| Brazil | 10 (5 MRSA / 5 MSSA) | 2 | 4 | 6 | 0 | 10 | 3 | 7 | 0 | 10 | 20 |
|  | 10 (≠ 5/5) | 1 | 0 | 0 | 1 | 1 | 8 | 0 | 1 | 9 | 10 |
|  | <10 | 2 | 1 | 0 | 5 | 6 | 0 | 0 | 5 | 5 | 11 |
|  | Total | 5 | 5 | 6 | 6 | 17 | 11 | 7 | 6 | 24 | 41 |
| Paraguay | 10 (5 MRSA / 5 MSSA) | 4 | 9 | 11 | 0 | 20 | 9 | 9 | 2 | 20 | 40 |
|  | 10 (≠ 5/5) | 1 | 5 | 0 | 1 | 6 | 2 | 1 | 1 | 4 | 10 |
|  | <10 | 4 | 4 | 1 | 0 | 5 | 8 | 1 | 2 | 11 | 16 |
|  | Total | 9 | 18 | 12 | 1 | 31 | 19 | 11 | 5 | 35 | 66 |
| Uruguay | 10 (5 MRSA / 5 MSSA) | 1 | 2 | 3 | 0 | 5 | 2 | 3 | 0 | 5 | 10 |
|  | <10 | 7 | 3 | 3 | 0 | 6 | 10 | 16 | 0 | 26 | 32 |
|  | Total | 8 | 5 | 6 | 0 | 11 | 12 | 19 | 0 | 31 | 42 |
| Total |  | 58 | 96 | 78 | 8 | 182 | 123 | 126 | 12 | 261 | 443 |

**Suppl Fig. 1**. Sampling map of StaphNET-SA study surveillance sites. Coloured pie charts depict the distribution of MRSA (red) and MSSA (light blue); pie size is scaled according to the number of isolates collected at each site.

*S. aureus* identification and antibiotic susceptibility tests (AST) were done in each centre by conventional methods (biochemical tests, disk diffusion, Vitek2, Phoenix, and/or Microscan). Antibiotics tested included: oxacillin (OXA), cefoxitin (FOX), erythromycin (ERY), clindamycin (CLI), gentamicin (GEN), ciprofloxacin (CIP), levofloxacin (LVX), rifampin (RIF), trimethoprim-sulfamethoxazole (SXT), minocycline (MNO), vancomycin (VAN), and linezolid (LZD). AST (interpreted as per CLSI 2019 guidelines [18]), origin of infection (community acquired (CA) or hospital acquired (HA), as per GLASS 2017 definition [19]), and other relevant demographic and epidemiological metadata (patient age and sex, date of isolation, geographic location, and gateway of bacteremia) were collected for each isolate using Epicollect5 (https://five.epicollect.net/) [20]. All isolates were sent to the School of Pharmacy and Biochemistry (FFyB-UBA), where species identification was confirmed by MALDI-TOF-MS. Antibiotic susceptibility was confirmed by the disk diffusion method as per CLSI 2019 guidelines. All *S. aureus* isolates were retested for methicillin susceptibility while susceptibility to other antibiotics was retested only when genomic results were discordant with AST results.

Multiple drug resistance (MDR) was defined as previously described [21] with the following modifications: i) susceptibility profiles for LVX and/or CIP were used to define fluoroquinolone resistance; ii) tetracycline (MNO), glycopeptide (VAN) and oxazolidinone (LZD) families were not included in the MDR definition due to the high number of isolates non tested for those antibiotics.

### Whole genome sequencing

Genomic DNA was extracted using the Qiacube system (QIAGEN), with the addition of Lysostaphin. DNA was quantified with the Quantus™ Fluorometer (Promega). Whole genome sequencing was performed at the Wellcome Sanger Institute on the Illumina HiSeq platform with 150-bp paired-end reads.

### QC, assembly and annotation

Genomes were quality controlled and assembled from short-read data with the GHRU-AMR pipeline [22] which is described in detail here: https://www.protocols.io/view/ghru-genomic-surveillance-of-antimicrobial-resista-bpn6mmhe. Default parameters were used for all software unless otherwise specified. Genomes were excluded from the study if more than 5% of the reads belonged to another species, or based on the quality of their assemblies if they contained more than 400 contigs, more than 10,000 ambiguous bases (Ns), their N50 was < 14,000, or their total length was at least 10% smaller than the smallest genome or at least 10% larger than the largest complete *S. aureus* genome in RefSeq, or their GC content was smaller than 32.4 or larger than 35.1 (based on the complete *S. aureus* genomes in RefSeq).

### In silico genotyping

The *spa* types were derived from assemblies using spaTyper (http://spatyper.fortinbras.us). MLST was determined from reads using the GHRU-AMR pipeline [22]. We used multiple approaches to determine the SCC*mec* type. First, we mapped the sequence reads to a SCC*mec* database [http://www.sccmec.org] of genes defining the *ccr* complex, the *mec* complex, and the J1 region [23] with ARIBA v2.14.6 [24], and determined the type and subtype based on the matches and Kondo typing scheme [23]. We complemented the SCC*mec* database with the genes from types XII and XIII. We also analysed the genomes with Staphopia [25], which yielded concordant results for 354/404 genomes. Assemblies of the remaining 50 genomes were analysed with SCCmecFinder [26], which determined a type/subtype for 39 additional genomes. The SCC*mec* cassette of 11 genomes were non-typable.

### Detection of antimicrobial resistance and virulence determinants, and mobile genetic elements

Detection of antimicrobial resistance determinants, virulence genes and mobile genetic elements (MGEs) was carried out with ARIBA v2.14.6 [24] and relevant databases. AMR determinants were determined with the GHRU-AMR pipeline [22], using the NCBI database (downloaded 2021-02-17)[27] and the pointfinder database (downloaded 2021-02-17)[28]. Virulence genes were detected using the VFDB database (downloaded 2021-08-27)[29]. Plasmid replicons were detected using Plasmidfinder database (downloaded 2021-09-10)[30]. Phage integrase types, intSaPI types, insertion sequences, and ICEs were detected as described previously [15].

### Pangenome analysis

Assemblies were annotated with the implementation of Prokka [31] in Panaroo v1.2.0 [32], which was used to determine the pangenome of 404 isolates. SNPs were identified from the resulting core-genome alignment with snp-sites v2.5.1 [33] and were used to build a maximum-likelihood (ML) phylogenetic tree with RAxML v8.2.8 [34] using the GTR+GAMMA model and 500 bootstrap replicates. Pairwise SNP differences were calculated with pairsnp v0.0.7 [35].

### Global context, variant detection and phylogenetic analysis

Genomes belonging to the most prevalent clonal complexes in the study (CC30, CC5, CC8, and CC398) were contextualised with global public genomes **(Suppl. Table 1)**.

Short paired-end reads were simulated from assemblies for genomes without short-read data available. Contigs smaller than 1kb were removed from the assemblies with seqkit v0.10.1(0.10.1--1)[36], and the 100 bp paired-end reads were simulated with depth 50x and insert length 500 using pIRS v2.0.2 (2.0.2--pl5.22.0_1) [37].

Reads were then mapped to the corresponding reference genomes **(Suppl. Table 1)** using the GHRU snp_phylogeny pipeline [22]. SNPs were identified with snp-sites v2.5.1 [33] from the resulting whole-genome alignments, after excluding regions of mobile genetic elements (MGEs) [38] and recombination with Gubbins v3.0.0 [39], and were used to build a ML-phylogenetic tree with IQ-Tree v1.6.10 [40], with ModelFinder to determine the best-fit model [41]. Branch support was estimated with SH-aLRT and ultrafast bootstrap (1000 replicates each) [42]. The resulting tree was rooted using an outgroup genome **(Suppl. Table 1)** that was omitted from the figures. AMR and virulence determinants, SCC*mec* type, and MGE were detected for these global collections of genomes with the same methods as described above.

The Microreact web application [43] was used for the integrated visualisation of phylogenetic trees, geographic, temporal data and other associated epidemiological and genetic data.

### Statistical methods

*p*-values were calculated by Fisher’s exact test for categorical variables, and by Pearson’s chi-squared for estimating differences in MSSA:MRSA proportions using R [44]. Wilcoxon rank-sum test, Cohen’s d and Cliff’s Delta values were used to estimate differences in pairwise SNPs distances between genomes of the same or different country using R and the effsize package [44, 45]. A *p-value* < 0.05 was considered significant in all cases.

Comparison of estimates of diversity were calculated for the MRSA and the MSSA subpopulations in our study. The operational taxonomic unit (OTU) was the sequence type (ST) and novel STs were considered distinct OTUs. Observed and asymptotic estimates of diversity (OTU Richness, Shannon index and Simpson index) and their corresponding 95% bootstrap confidence interval were calculated with iNEXT v.2.0.20 [46] with parameters datatype = abundance, se = TRUE, conf = 0.95, nboot = 50, and endpoint = 500.

## Results

### Survey Summary

*S. aureus* isolates were collected from blood cultures in 58 hospitals from Argentina, Bolivia, Brazil, Paraguay, and Uruguay between April and October 2019. Thirty hospitals reached the objective of ten isolates, out of which 20 hospitals collected the target ratio of 1:1 MSSA:MRSA, while the remaining ten hospitals were unable to collect either five MRSA or five MSSA isolates during the sampling period and submitted a different MSSA:MRSA ratio. Twenty-eight hospitals were unable to collect 10 isolates **(Table 1)**.

Thus, a total of 443 isolates were collected and the proportion of MRSA and MSSA in our study was 41.1% (182/443) and 58.9% (261/443), respectively (**Table 1**). Notably, MRSA isolates were recovered mainly in the northeast of the sampled region (**Supplementary Fig. 1**).

*S. aureus* blood isolates were recovered predominantly from male (287/443, 64.7%) and adult patients (>18 years) (315/443, 71.1%). The gateway of bacteremia was mainly skin and soft tissue infections (SSTI) (105/443, 23.7%) followed by respiratory (86/443, 19.4%) and catheter (75/443, 16.9%). Infections with a catheter origin were more frequent in MSSA than in MRSA (Fisher’s exact test, *p* = 8.64E-05) (**Table 2**).

**Table 2.** Infection origin and gateway of bacteremia of MRSA and MSSA collected isolates. CA: Community Acquired. HA: Hospital Acquired. ND: Data not available.

| Gateway of bacteremia | *S. aureus*<br>(n%) | MRSA |  |  | Total MRSA<br>n(%) | MSSA |  |  | Total MSSA<br>n(%) |
| --- | --- | --- | --- | --- | --- | --- | --- | --- | --- |
|  |  | CA n(%) | HA n(%) | ND n(%) |  | CA n(%) | HA n(%) | ND n(%) |  |
| Skin and soft tissue infection | 105<br>(23.7%) | 42 (40%) | 12 (11.4%) | 0 (0%) | 54 (51.4%) | 36 (34.3%) | 14 (13.3%) | 1 (1.0%) | 51 (48.6%) |
| Respiratory | 86<br>(19.4%) | 21 (24.4%) | 14 (16.3%) | 0 (0%) | 35 (40.7%) | 23 (26.7%) | 26 (30.2%) | 2 (2.3%) | 51 (59.3%) |
| Catheter | 75<br>(16.9%) | 3 (4%) | 17 (22.7%) | 0 (0%) | 20 (26.7%) | 10 (13.3%) | 44 (58.7%) | 1 (1.3%) | 55 (73.3%) |
| Data not available | 54<br>(12.2%) | 7 (13.0%) | 7 (12.9%) | 6 (11.1%) | 20 (37.0%) | 14 (25.9%) | 13 (24.1%) | 7 (13.0%) | 34 (63.0%) |
| Other | 41 (9.3%) | 9 (22%) | 7 (17.1%) | 2 (4.9%) | 18 (43.9%) | 12 (29.3%) | 10 (24.4%) | 1 (2.4%) | 23 (56.1%) |
| Bone | 31 (7.0%) | 9 (29.0%) | 4 (12.9%) | 0 (0%) | 13 (41.9%) | 16 (51.6%) | 2 (6.5%) | 0 (0%) | 18 (58.1%) |
| Endovascular | 30 (6.8%) | 5 (16.7%) | 7 (23.3%) | 0 (0%) | 12 (40%) | 9 (30%) | 9 (30%) | 0 (0%) | 18 (60%) |
| Surgical wound | 21 (4.7%) | 0 (0%) | 10 (47.6%) | 0 (0%) | 10 (47.6%) | 3 (14.3%) | 8 (38.1%) | 0 (0%) | 11 (52.4%) |
| Total | 443<br>(100%) | 96<br>(21.7%) | 78<br>(17.6%) | 8 (1.8%) | 182 (41.1%) | 123 (27.8%) | 126<br>(28.4%) | 12 (2.7%) | 261<br>(58.9%) |

We determined the resistance rates for 12 antibiotics overall, and for combinations of MRSA or MSSA and HA or CA. The highest resistance rates overall were found for erythromycin (29.7%) and clindamycin (26.9%), followed by gentamicin (16.0%) and fluoroquinolones (ciprofloxacin 12.0% and levofloxacin 8.5%)(**Table 3**). All tested isolates were susceptible to vancomycin (n=296) and linezolid (n=196).

**Table 3.**
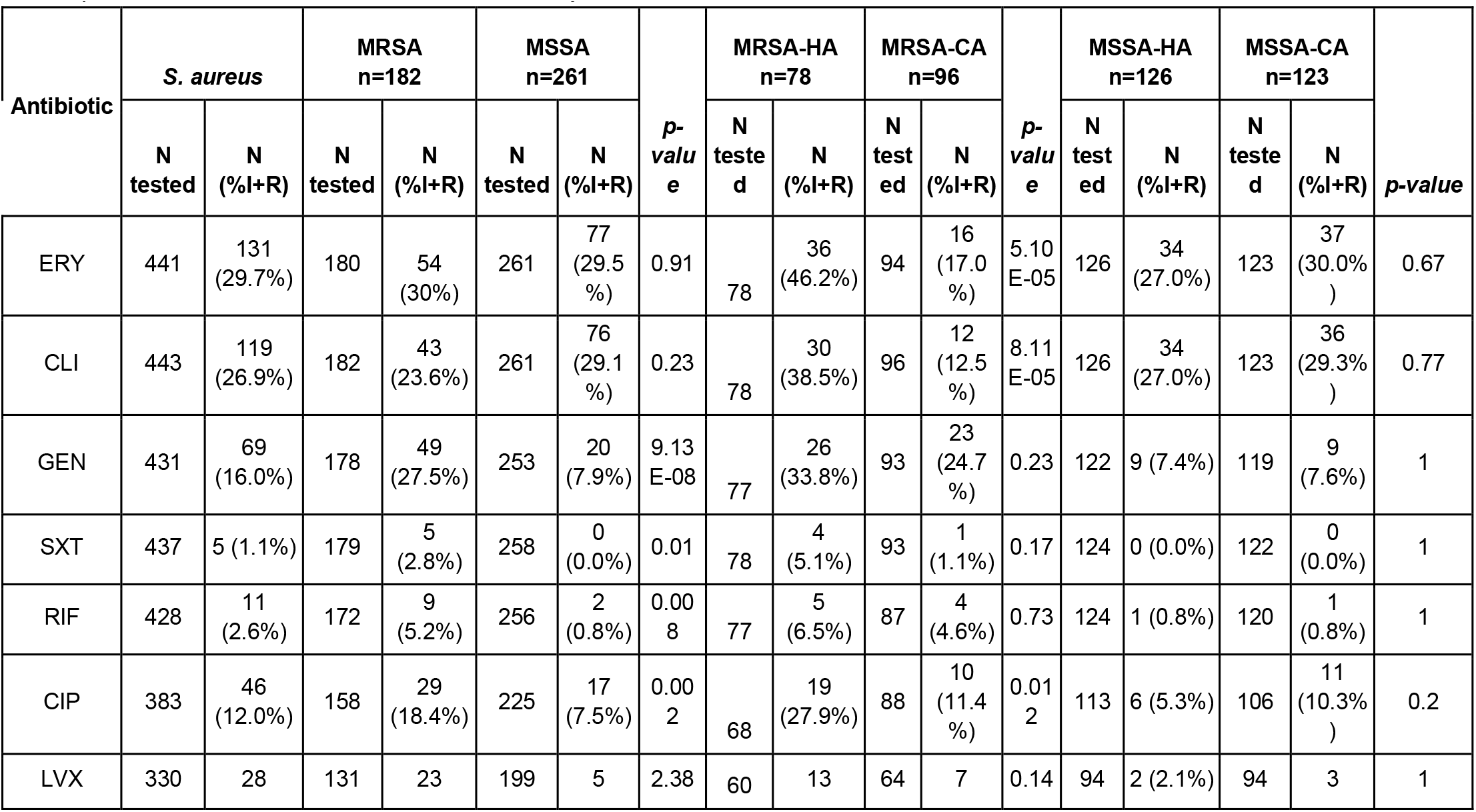
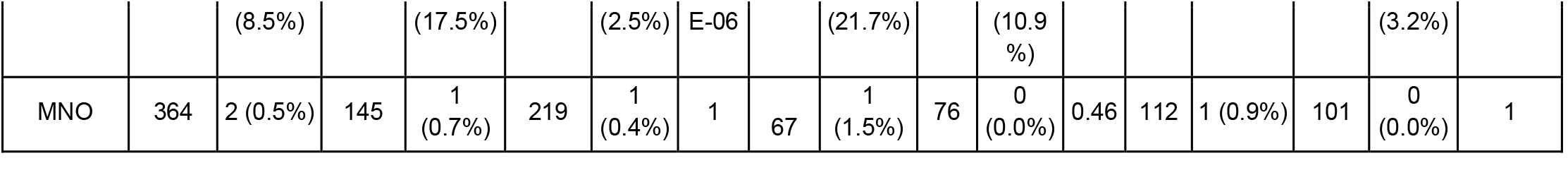
Antimicrobial susceptibility of *S. aureus* isolates. N tested=number of isolates tested for any given antibiotic. Results are expressed as percent of tested isolates that are non-susceptible (% Intermediate + % Resistance, %I+R). ERY: Erythromycin, CLI: Clindamycin, GEN: Gentamicin, SXT: Trimethoprim-Sulfamethoxazole, RIF: Rifampin, CIP: Ciprofloxacin, LVX: Levofloxacin, MNO: Minocyclin.

However, MRSA strains exhibited higher antimicrobial resistance rates for fluoroquinolones (ciprofloxacin and levofloxacin), rifampin, trimethoprim-sulfamethoxazole and gentamicin than MSSA (Fisher’s exact test, *p<0*.*05*, **Table 3**). A comparison of the origin of the infection showed that HA-MRSA resistance rates to ciprofloxacin, erythromycin, and clindamycin were significantly higher than those of CA-MRSA (Fisher’s exact test, *p<0*.*05*, **Table 3**). No significant differences in resistance rates were found between CA-MSSA and HA-MSSA (**Table 3**). Nevertheless, differences in these proportions were found between countries, like Bolivia, where erythromycin and clindamycin resistance are not so prevalent in MSSA (11.8% and 14.7%, respectively) **(Suppl. Table 2)**.

### Different populations of MRSA and MSSA in South America

A total of 404 out of 443 collected *S. aureus* isolates in 58 hospitals passed all quality controls (**Suppl Fig. 2**), 239 of which were defined as MSSA and 165 as MRSA based on the absence or presence of the *mecA* gene, respectively.

**Supplementary Fig. 2**. Diagram flow showing confirmation and quality control performed on *S. aureus* isolates and genomes in this study.

Among them, we identified 59 different allelic profiles/sequence types (STs) grouped into 14 clonal complexes (CCs) and 9 singletons. The MSSA were more diverse than the MRSA (as measured by the Shannon and Simpson diversity indexes, **Suppl. Table 3**), with 44 STs grouped into 14 CCs and 8 singletons, and 20 STs grouped into 7 CCs and 2 singletons, respectively. CC30 (18.6%), CC5 (17.6%), and CC8 (15.8%) and CC398 (11.1%) were the most prevalent overall and recovered from all five countries (**Suppl Fig. 3, Figure 1**). Eighty-seven percent of the MRSA belonged to three CCs, CC30 (34.5%), CC5 (30.3%), and CC8 (22.4%), while CC398 (18.8%), CC8 (11.3%) and CC1 (9.6%) were the most prevalent CCs among the MSSA population (39.7%, **Figure 1A, Suppl Fig. 3**).

**Figure 1.**
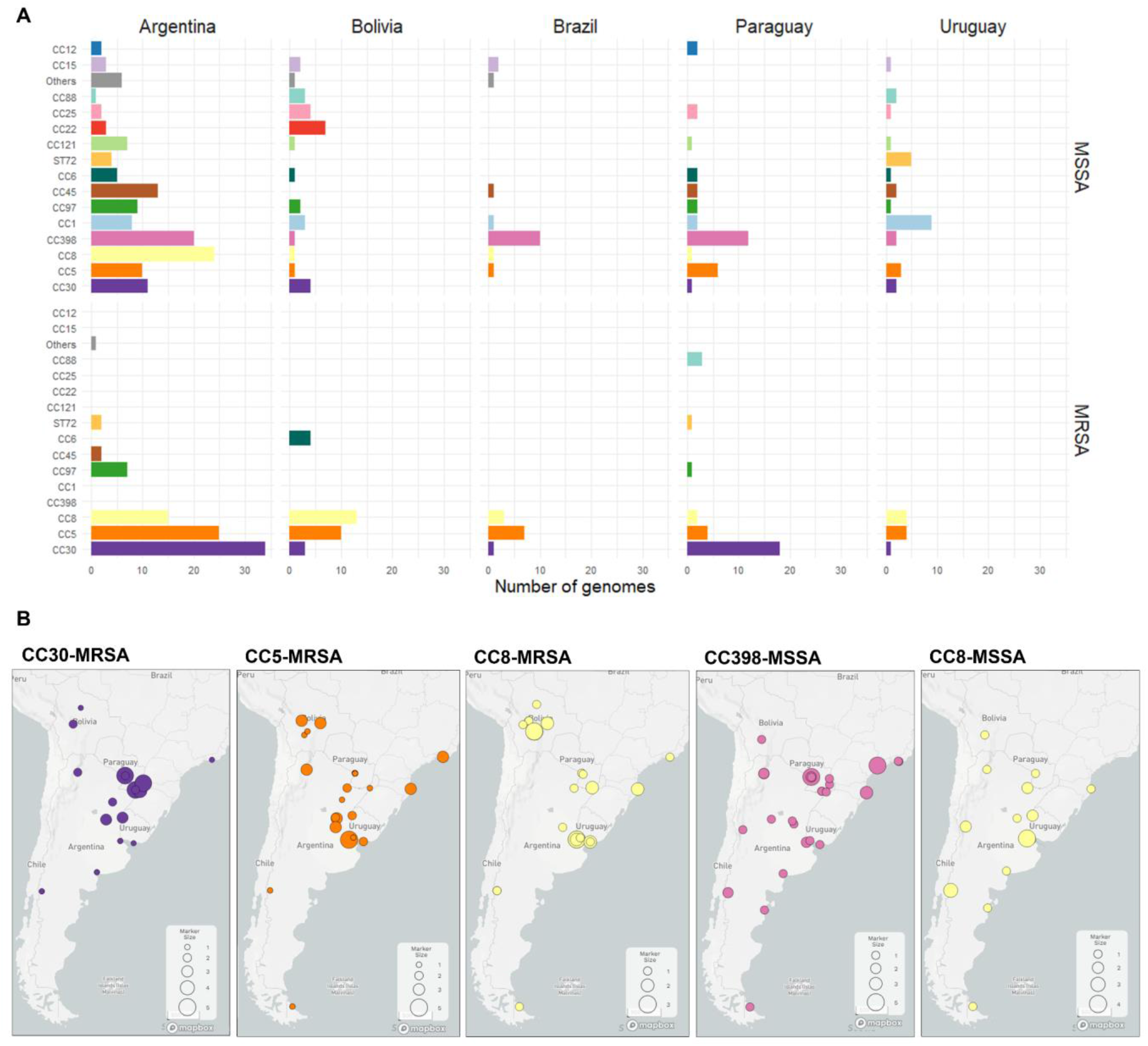
Distribution of clonal complexes by country. **A)** Frequency of CCs by country and MRSA or MSSA. STs comprising less than 3 genomes are grouped under “Others”. **B)** Geolocalization of major clonal complexes in the region. The pie charts on the maps depict the distribution of CCs at each sampling site; the pie size is scaled by the number of genomes collected at each site.

**Supplementary Fig. 3**. Distribution of clonal complexes in the 404 *S. aureus* genomes by MRSA or MSSA. STs comprising less than 3 genomes are grouped under “Others”. Bars are coloured as described in the legend.

Notably, CC398-MSSA is not only described here for the first time as the most prevalent MSSA lineage but is also widely disseminated throughout the region (**Figure 1B**). CC398-MSSA was found to be the dominant MSSA lineage in Brazil and Paraguay, and the second most prevalent in Argentina, where CC8-MSSA is also widely distributed. CC398-MSSA harbours exclusively the *ermT* gene, mainly *spa* type *t1451* and lacks *lukF/S-PV* genes (**Figure 2**).

**Figure 2.**
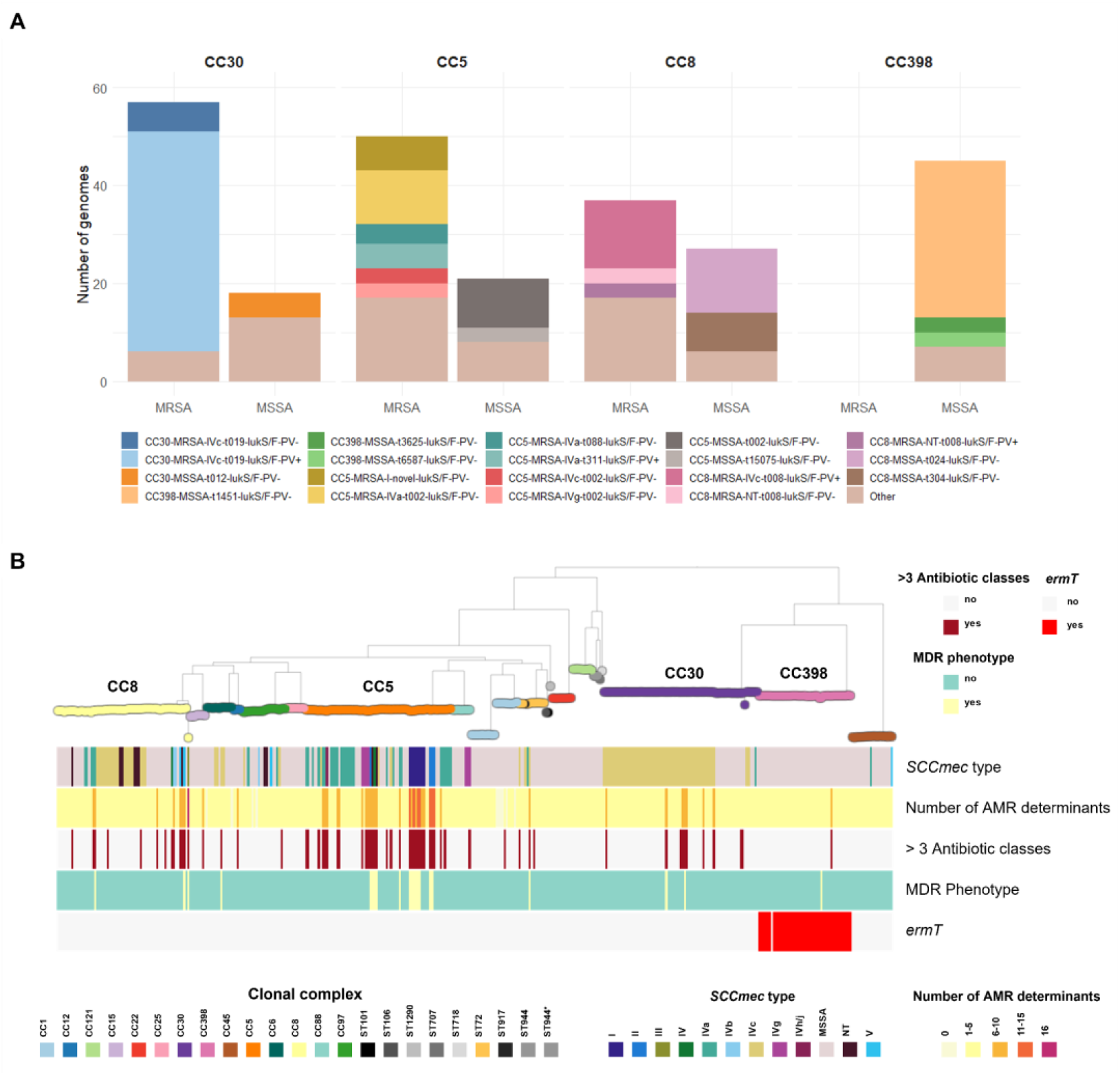
**A)** Genotypes of the main clonal complexes in the study. “Other” include genotypes present in less than 3 genomes. **B)** Maximum Likelihood tree of 404 genomes inferred from 156868 SNP sites identified on 2182 core genes (Panaroo) with RAxML. Midpoint rooted. 500 bootstrap replicates. Tree nodes and blocks are coloured as described in the legend. MDR phenotype was defined as previously described (Magiorakos 2012) for all antibiotics tested including: gentamicin, cefoxitin, erythromycin, clindamycin, fluoroquinolones, minocycline, trimethoprim-sulfamethoxazole, rifampin, vancomycin and linezolid. “>3 antibiotic classes”: genome with AMR determinants for >3 antibiotic classes including the ones detailed above plus phenicols, fusidic acid, fosfomycin, mupirocin and bleomycin (not tested phenotypically).

Nonetheless, differences in the frequency of clonal complexes were also found amongst countries, despite few similarities between those sharing borders (**Figure 1**). Less prevalent MSSA clonal complexes overall still prevail in Uruguay (CC1-MSSA) and Bolivia (CC22-MSSA). CC30-MRSA remained prevalent in Argentina and Paraguay consistent with previous reports [47–49], while CC8-MRSA and CC5-MRSA prevailed in Bolivia and Brazil, respectively (**Figure 1A**).

The MSSA and MRSA subpopulations of CC30, CC5, and CC8 could be differentiated into genotypes defined by molecular markers traditionally used to describe the epidemiology of *S. aureus*, i.e., the SCC*mec* type, *spa* type, and the presence of *lukF/S*-PV genes (**Figure 2**).

Genotypes CC30-MRSA-IVc-t019-*lukF/S*-PV*+*, CC8-MRSA-IVc-t008-*lukF/S*-PV*+ and* CC5-MRSA-IV*-t002-lukF/S-*PV*-* (carrying different *SCCmec* IV subtypes) were the more prevalent in the MRSA subpopulation (**Figure 2**). Some *spa* types (such as *t311* and *t002)* were found in both MRSA and MSSA within the same clonal complex (like CC5), indicating that the SCC*mec* cassette could have been acquired/lost during the evolution of this CC in the South American cone. However, *spa* types *t019 and t008*, found only in CC30-MRSA and CC8-MRSA, respectively, have no counterpart in MSSA (**Figure 2A**), suggesting that the SCC*mec* cassette was acquired prior to their arrival in the region.

In agreement with AST results, *S. aureus* genomes in our collection usually carry a relatively low number of AMR determinants (360/404, 80.1% carry 5 or less AMR determinants). 21/404 (5.2%) isolates in our collection were phenotypically MDR, i.e., resistant to at least 3 antibiotic classes. However, we found 64/404 (15.8%) genomes with AMR determinants to at least 3 antibiotic classes, due to the *in-silico* detection of resistance to antibiotics not routinely tested, such as fosfomycin (*fosB*, 245/404). Nonetheless, the concordance between genotypic and phenotypic resistance was high (>98.5%) for eight antibiotics analysed **(Suppl. Table 4**). MDR isolates (21/404) mostly belonged to minor genotypes within CC5, all of them *lukF/S-PV negative* (ST5-MRSA-I-*tnovel*|*others*, ST105-MRSA-II-*t002*|*t985*, ST100-MRSA-NT|IV-*t002*, and ST5-MRSA-II-*t509*) (**Figure 2B**). Different genotypes within CC5 contributed to the higher AMR rates in MRSA (fluoroquinolones, gentamicin) and in HA-MRSA (fluoroquinolones and MLSb). CC30-MRSA-IVc*-t019* is also responsible for gentamicin resistance rates while trimethoprim-sulfamethoxazole resistance was linked to CC6-MRSA-IVc-*t701* (**Figure 2B, Suppl. Fig. 4**).

**Suppl Fig.4**. AMR determinants and AMR phenotypes. Maximum Likelihood tree of 404 genomes inferred from 156868 SNP sites identified on 2182 core genes (Panaroo) with RAxML. Midpoint rooted. 500 bootstrap replicates. Tree nodes and blocks are coloured as described in the legend.

### Prevalent lineages in a global context

To investigate the evolutionary relationships between the South American MRSA and MSSA, we contextualised genomes belonging to CC30, CC5, CC8 and CC398 with published genomes from different countries and continents, which we describe next.

### CC30

Several epidemic clones have emerged within CC30, i.e. MSSA-PVL+ phage type 80/81 (ST30 [50]), hospital-associated EMRSA-16 (ST36 [51]), MRSA-PVL+ South West Pacific (SWP) clone (ST30, [52, 53], and EMSSA-ST30 [6]. A prospective study of *S. aureus* bloodstream infections from nine South American countries showed that MRSA-ST30 was prevalent in Argentina but less so in other countries [8]. The Argentinean MRSA-ST30 were subsequently shown to be diverse, with most isolates belonging to one geographically disseminated clade (ARG-4) related to the SWP clone, but differing from it by the head of the PVL phage [15].

The CC30 genomes from this study (n=75) were mostly found within the MRSA-ST30 (n=54) and the EMSSA-ST30 (n=13) clones, and closely related to public South American and European genomes. One genome from Argentina and 1 from Bolivia clustered with the epidemic phage type 80/81 reference genome 55_2052 (**Figure 3A**) and within the more divergent ST34, respectively.

**Figure 3.**
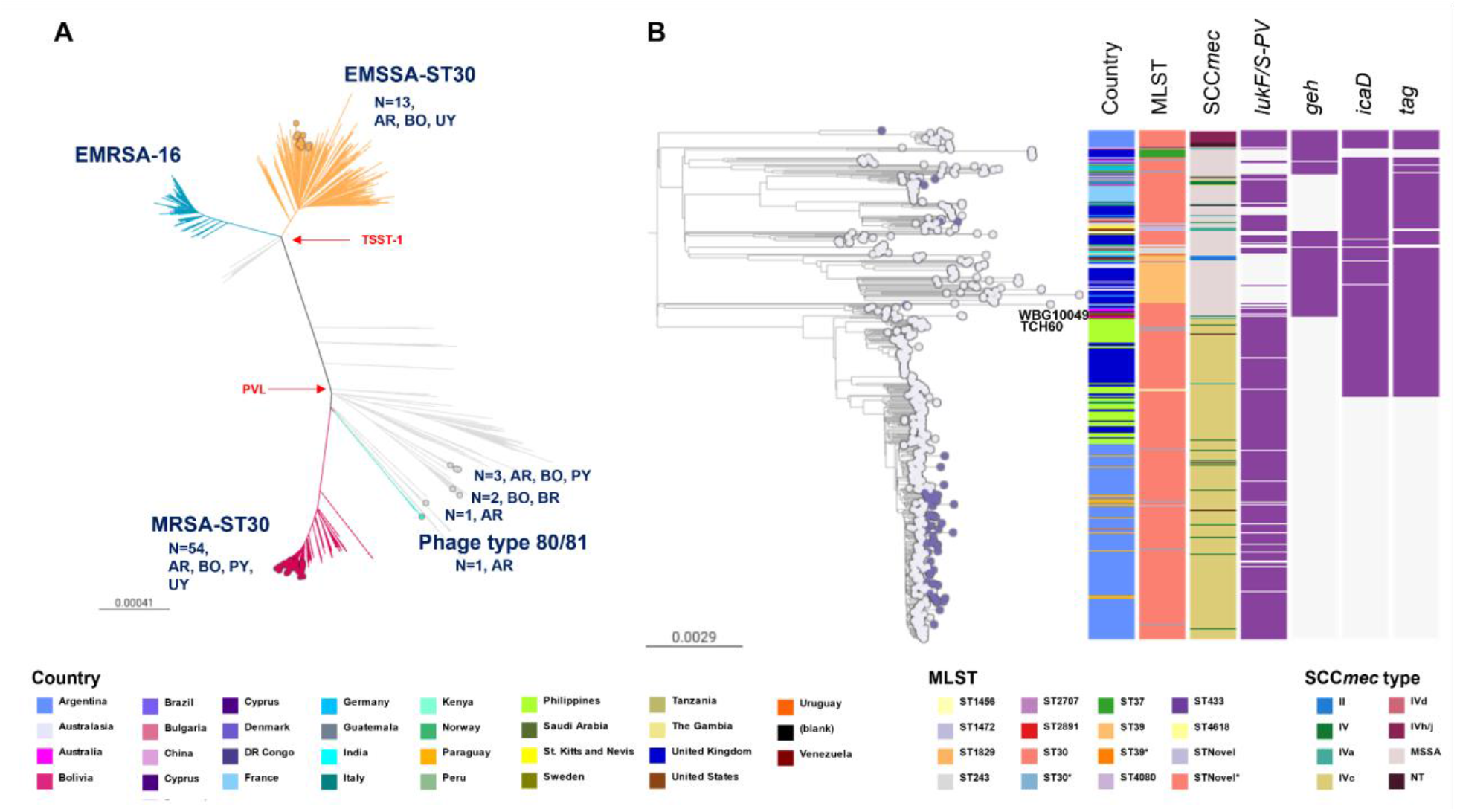
**A)** CC30 global phylogeny (the divergent ST34 clade is not represented). Branches are coloured by clade: EMSSA-ST30 (orange), EMRSA-16 (blue), MRSA-ST30 (red), phage type 80/81 (aqua). Coloured nodes represent StaphNET-SA genomes, detailing number and country of origin. AR: Argentina, BO: Bolivia, BR: Brazil, PY: Paraguay, UY: Uruguay. Project is available in microreact https://microreact.org/project/cc30-global-context **B)** Detailed ST30 phylogeny including 61 genomes from our collection. Outgroup-rooted phylogenetic tree inferred from 61,666 SNP sites obtained after mapping the genomes to the complete genome of strain TCH60 and masking regions of recombination and MGEs. Leaf nodes are coloured by collection: StaphNET-SA (violet), global context (grey). Coloured blocks represent the presence of an intact genetic determinant (purple) or feature as described in the legend. For both trees, the scale bars represent the number of single nucleotide polymorphisms (SNPs) per variable site. Project is available in microreact https://microreact.org/project/emrsa-st30-global-context

As most CC30 genomes from this study (61/75) were found within a clade of 598 genomes supported by a 100% bootstrap value and including the MRSA-ST30 clone, we inferred a more detailed phylogenetic tree of this group (**Figure 3B**). The tree revealed multiple events of acquisition/loss of type IV and type II SCC*mec* cassettes. The notable presence of one monophyletic MRSA clade (100% bootstrap support) comprising 381 genomes from 10 countries with type IVc SCC*mec* was indicative of substantial clonal expansion and global dissemination. The basal branches include two contemporary genomes from Venezuela on a long branch, and a cluster of three genomes from Australia and USA representing the SWP clone (1999-2004). The isolates from the southern cone of South America, including those from this study (54/75, mainly of CA origin), form a single clade comprising 229 genomes (100% bootstrap support), consistent with one major introduction followed by clonal spread and geographic dissemination. The basal genomes in the South American clade are from Buenos Aires, Argentina, and genomes from Paraguay (n=18), Bolivia (n=18), Uruguay (n=1) and UK (n=1) are interspersed with genomes from Argentina (n=207). Notably, genomes from 2019 (this study) are found on longer branches than those from 2005-2014 within this South American clade, and shared *geh, icaD, and tag* mutations already described in the Argentinean genomes [15] **(Figure 3B)**.

### CC5

The diverse set of CC5 genotypes from our collection, predominantly MRSA (50/71; 70.4%) **(Figure 2A)**, grouped within the four previously defined clades that have caused infections globally [6, 54]: CC5-Basal, CC5-I, CC5-II A, and CC5-II B **(Figure 4)**.

**Figure 4.**
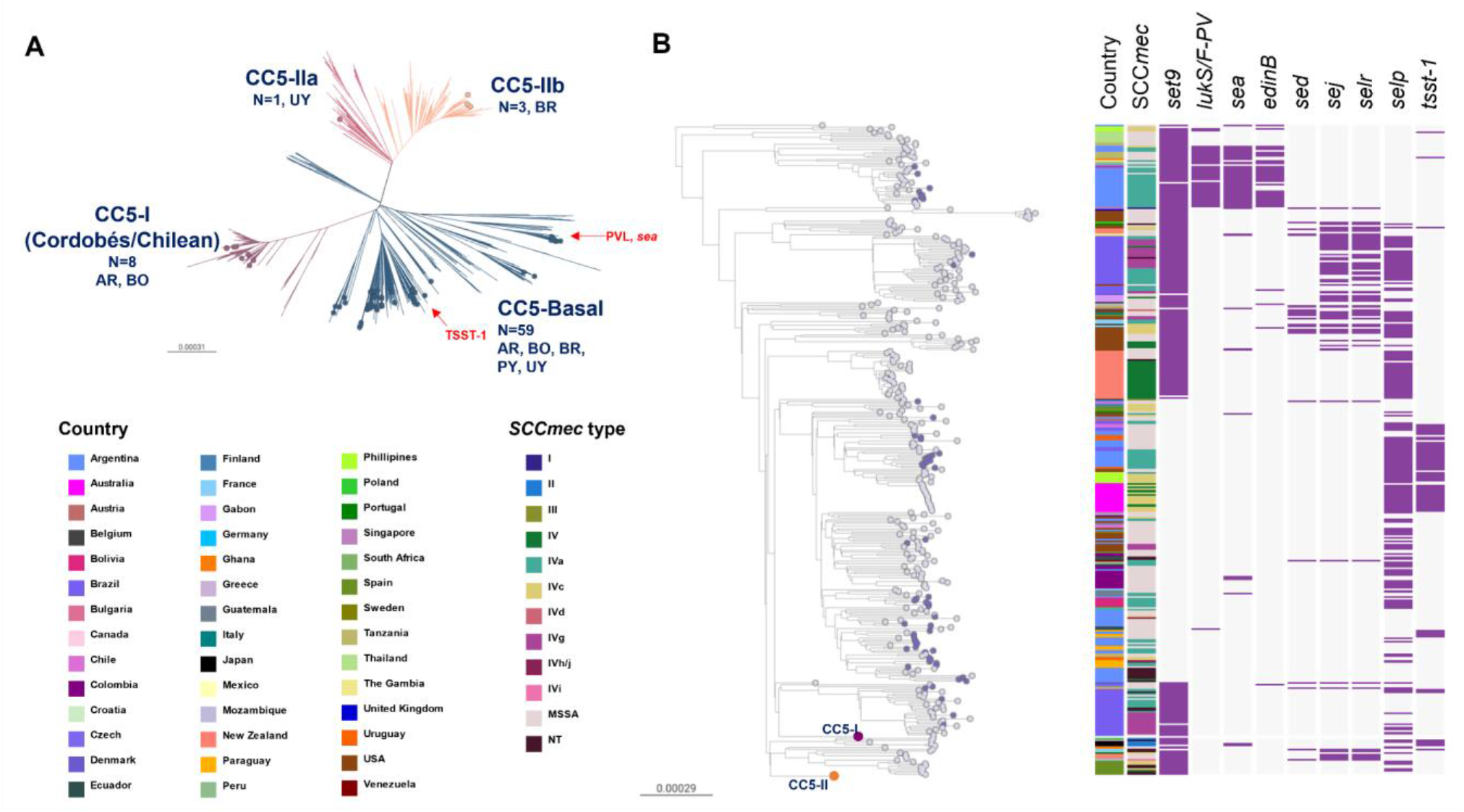
**A)** CC5 global phylogeny. Outgroup-rooted phylogenetic tree inferred from 66837 SNP sites obtained after mapping the genomes to the complete genome of strain JH1 (ST105) and masking regions of recombination and MGEs. Branches are coloured by clade: CC5-Basal (blue), CC5-I (violet), CC5-II-A (light violet), CC5-II-B (salmon). AR: Argentina, BO: Bolivia, BR: Brazil, PY: Paraguay, UY: Uruguay. **B)** CC5-Basal clades. Same tree from the phylogeny in Figure A but CC5-I and CC5-II clades are collapsed as a purple (263 genomes) and salmon (436 genomes) circles, respectively. Leaf nodes are coloured by collection: global context (grey), StaphNET-SA (violet). Coloured blocks represent the presence of an intact genetic determinant: virulence gene (violet). Country and *SCCmec* type colours are described in the legend. For both trees, the scale bars represent the number of single nucleotide polymorphisms (SNPs) per variable site.Project is available in microreact https://microreact.org/project/cc5-global-context

However, the CC5-MRSA lineages once predominant in the region both in the community (ST5-MRSA-IV-*t311* with *lukS/F-PV* and *sea* genes) and in hospitals (CC5-I: ST5-MRSA-I; CC5-II: ST105-MRSA-II and ST5-MRSA-II) [8, 47, 55–57] were scarcely represented and instead replaced by representatives of the CC5-Basal clades (59/71, MSSA or MRSA-IV with *spa* types *t002, t311* and related) which are typically not MDR, frequently lack the *set9/ssl4* gene (44/59 genomes), and harbour other toxin genes in a variable manner (**Figure 4B**). On the other hand, the persisting members of CC5-I (8/71) and CC5-II (4/71) contribute to CC5 showing the highest AMR rates of any CC in our study (**Figure 2**).

Our CC5-Basal genomes are polyphyletic and are interspersed with genomes from South America and also other continents (**Figure 4B**), although the global contextualization revealed clades specific to Brazil and Argentina that were not obvious from our data alone **(Figure 4B)**. Multiple events of acquisition/loss of SCC*mec* IV subtypes (IVa, IVc, IVg, IVh/j) were also evident from the tree.

Although CC5-MRSA-IV-*t002* were initially described in the community in Argentina and Brazil [47, 58], they were recovered from both CA (6/59) and HA (10/59) origins in our study. In parallel, we found MDR genomes recovered from CA origin within successful HA-MRSA clades among the less prevalent strains (ST5-MRSA-II and ST105-MRSA-II in Brazil and Uruguay), reinforcing the already described blurred boundaries between the community and the hospital.

### CC8

While the relevance of CC8 in South America has been previously linked to the epidemic spread of MRSA lineages (Brazilian clone and USA300-SAE) [10, 55], we found a substantial prevalence of MSSA within this clonal complex (27/64, 42.2%). The majority of our genomes grouped within the previously described USA300-Early Branching (28/64) and USA300-SAE (24/64, MRSA-IVc-*t008*-*lukF/S-PV*+-COMER+) clades (**Figure 5**).

**Figure 5.**
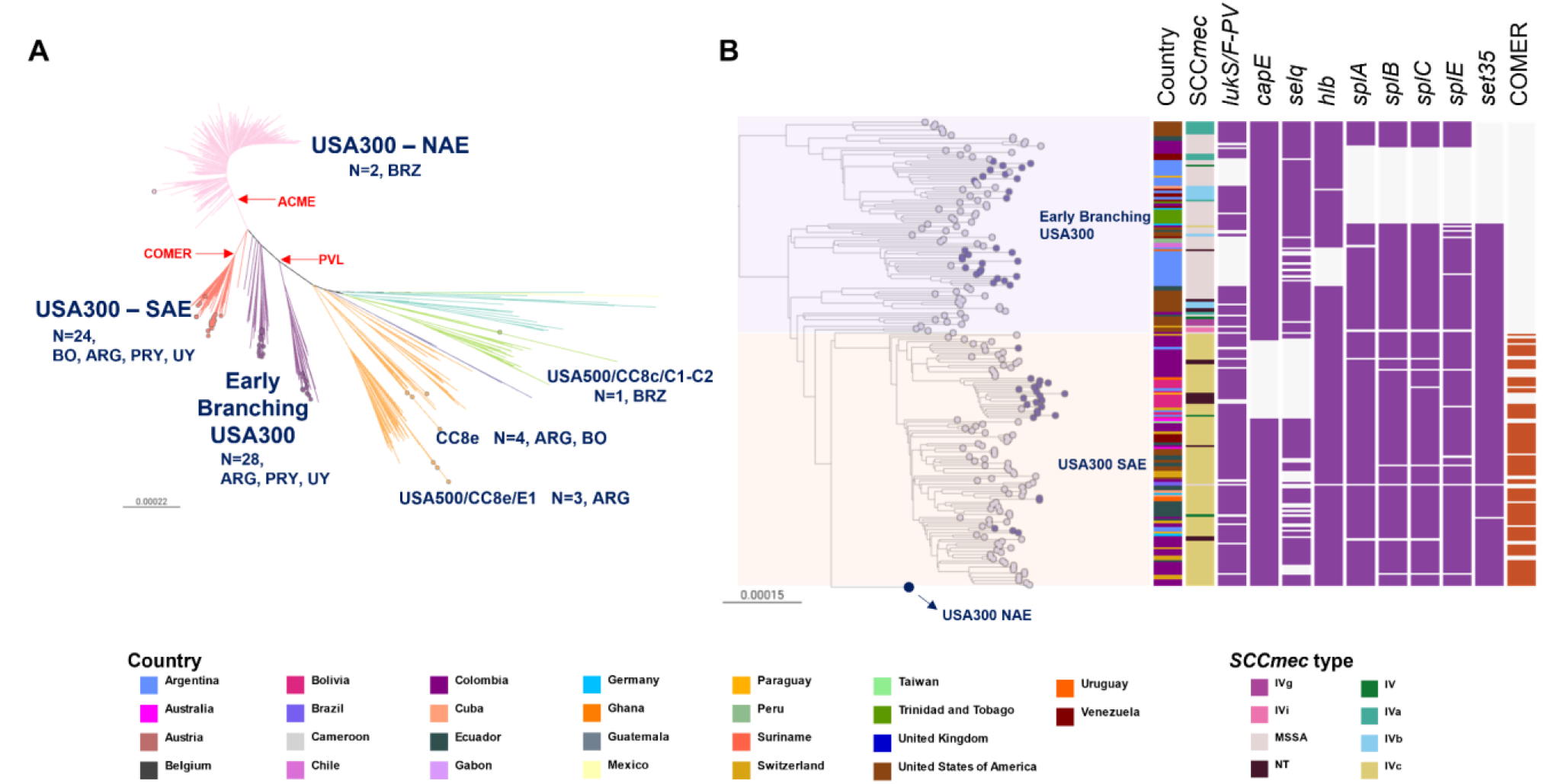
**A)** CC8 global phylogeny (more divergent Iberian and ST239 clades are not represented). Outgroup-rooted phylogenetic tree inferred from 66899 SNP sites obtained after mapping the genomes to the complete genome of strain USA300-FPR3757 (ST8) and masking regions of recombination and MGEs. Coloured nodes represent StaphNET-SA genomes, detailing number, and country of origin. AR: Argentina, BO: Bolivia, BR: Brazil, PY: Paraguay, UY: Uruguay. **B)** Detail of the USA300 subtree from A.The USA300-NAE clade (760 genomes) is collapsed as a blue circle. Leaf nodes are coloured by collection:StaphNET-SA (violet), global context (grey). Coloured blocks represent the presence of an intact genetic determinant: virulence gene (violet), COMER (brown). Country and SCC*mec* type colours are described in the legend. For both trees, the outgroup is omitted, and scale bars represent the number of single nucleotide polymorphisms (SNPs) per variable site. Project is available in microreact https://microreact.org/project/cc8-global-context

We only found a few genomes related to other CC8 lineages [10, 59, 60] circulating in Brazil (Iberian/CC8b n=1, USA500/CC8c n=1, and USA300 NAE n=2), Bolivia (CC8e n=1), and Argentina (CC8e n=6, and ST239 Brazilian clone n=1).

The USA300-Early Branching genomes were predominantly MSSA widely spread in Argentina (24/28) without a monophyletic origin, suggesting independent introductions (**Figure 1, Figure 5B**). One of the early branches comprised 11 genomes of CA origin (7/11) with *spa* type *t304* (8/11) from the centre and north of Argentina (10/11) and Paraguay (1/11) (**Suppl Fig. 5A**). The other early branch comprised 17 genomes of HA origin (13/17) with *spa t024* (11/17) in the centre and south of Argentina (16/17) and Uruguay (1/17), showing the sporadic acquisition of *erm*C (**Suppl Fig. 5B**).

Surprisingly, like related USA300-Early branching genomes from South America (2007-2013), most of our genomes lacked *lukS/F-PV* and other virulence genes (**Figure 5B, Suppl Fig. 5**) [10, 59].

**Suppl Fig. 5**. Geographic distribution of USA300 Early Branching clades. Leaf nodes are coloured by Country. Coloured blocks represent the presence of an intact genetic determinant: virulence gene (violet). Country and SCC*mec* type colours are described in the legend. For both trees, the outgroup is omitted, and scale bars represent the number of single nucleotide polymorphisms (SNPs) per variable site.

The MRSA USA300-SAE lineage known to be prevalent in the north of South America [8, 10] appears to have been introduced into our region on several occasions (**Figure 5B**). One such introduction comprised most of our USA300-SAE genomes (18/24) forming a distinct monophyletic group (100% bootstrap support) widely disseminated across Bolivia (14/18), consistent with the clonal expansion of this lineage in this country and multiple transmission events to Argentina, Paraguay, Uruguay and Switzerland. Of note, the cluster from Bolivia has a clear CA origin (13/18), and lacks functional *selq* and *capE* genes (**Figure 5B**).

### CC398

CC398 was originally described as a livestock-associated lineage, but it is also capable of causing human-to-human transmitted infections as reported from several countries. Phylogenetic evidence later revealed the existence of two distinct CC398 clades (livestock- and human-associated) differing in their repertoire of mobile genetic elements and virulence genes [61–65].

The CC398 genomes from this study were recovered from both CA (26/45) and HA (15/45) origin and belonged to *spa* types *t1451* and *t571*, typically linked to human origin (**Figure 1**) [65, 66]. Our phylogenetic analysis positioned them within the human-associated clade, closely related to both clinical and carriage isolates (**Figure 6**) and lacking *mecA* and *Tn916* (associated with *tetM*) but carrying the *Sa3int* phage-encoded Immune evasion cluster (IEC) genes *chp* and *scn* (but not *sak)* (**Figure 6B**).

**Figure 6.**
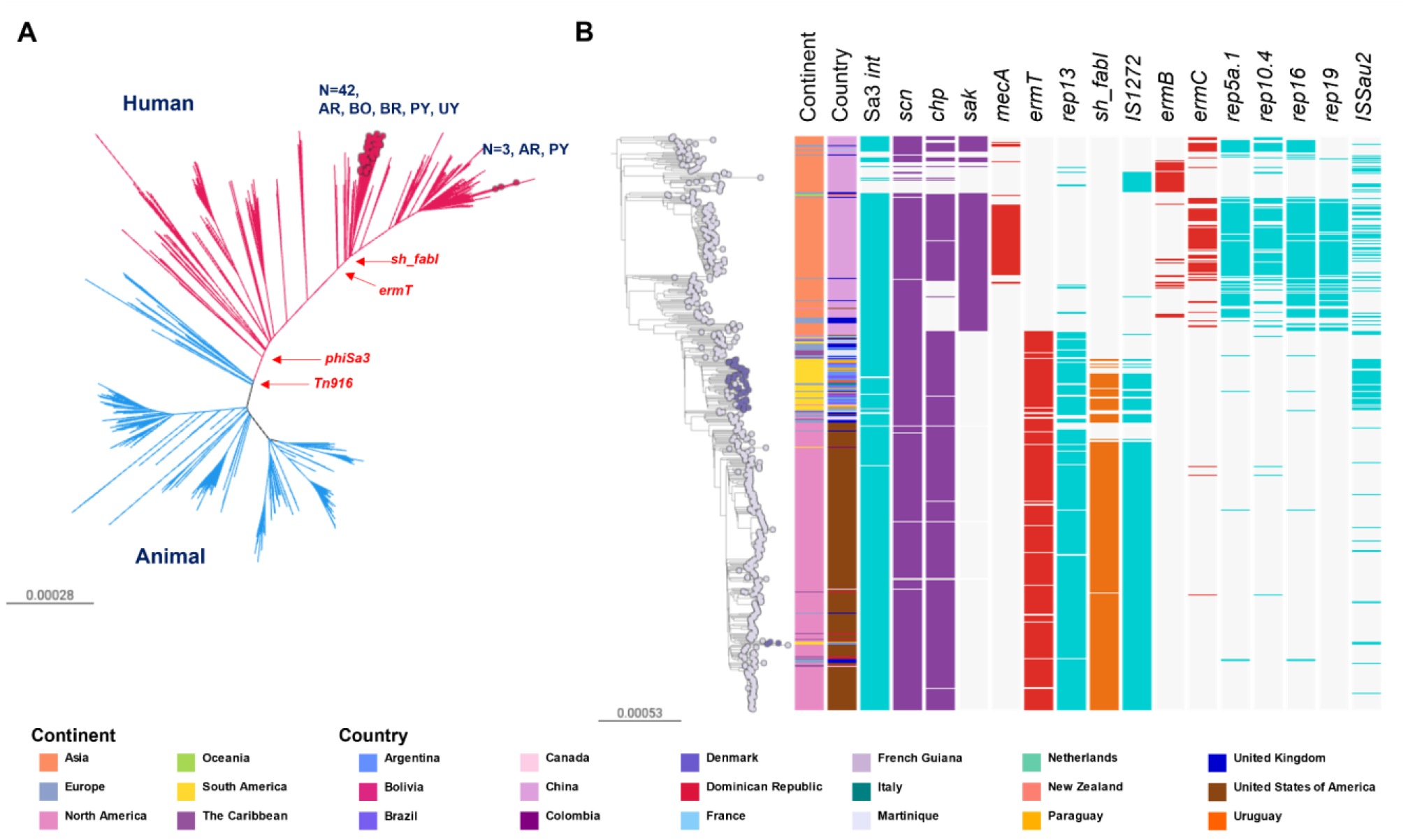
**A)** CC398 global phylogeny. Outgroup-rooted phylogenetic tree inferred from 71097 SNP sites obtained after mapping the genomes to the complete genome of strain S0385 (ST398) and masking regions of recombination and MGEs. Branches are coloured by the host association, animal (blue), human (red). Coloured nodes represent StaphNET-SA genomes, detailing number and country of origin. AR: Argentina, BO: Bolivia, BR: Brazil, PY: Paraguay, UY: Uruguay. **B)** Human-associated CC398 subtree from A. Leaf nodes are coloured by collection: StaphNET-SA (violet), global context (grey). Coloured blocks represent the presence of an intact genetic determinant: AMR gene (red), virulence gene (violet), mobile genetic element (aqua), triclosan resistance gene (orange). Country and continent colours are described in the legend. For both trees, scale bars represent the number of single nucleotide polymorphisms (SNPs) per variable site. Project is available in microreact https://microreact.org/project/cc398-global-context

Nested epidemics could be distinguished within the human-associated clade, one localised in China with occasional transmission to other countries (mainly the UK); a second one in South America involving the majority of the isolates from this study (42/45, from all 5 countries) clustering as a well-supported group (100% bootstrap) with ancestral relationships to genomes from the Caribbean, the USA, Europe and China; and a third epidemic mainly localised in the USA that sporadically seeded the Caribbean and South America, including three genomes from this study (2 Argentina, and 1 from Paraguay) (**Figure 6B**).

The majority of the genomes in the South American epidemic carried the *ermT* gene (generally found on the same assembly contig as the *rep13* replicon sequence), the insertion sequences *ISSau1/IS431* and the *cadDX* operon. Manual inspection of assembled genomes suggested that these plasmidic genes have a chromosomal location [65, 67, 68]. We also found *IS1271* and *sh-fabI* (associated with triclosan resistance [69]) in 33/45 genomes. The presence of *ermT, rep13, IS1272 and sh-fabI*, was also characteristic of the epidemic in the USA, but not of that in China, where the genomes instead carried *ermB* or *ermC* and other associated plasmidic replicons. On the other hand, an intact copy of *ISSau2*, prevalent in genomes from our collection and in public genomes from Brazil and Italy found within the South American epidemic (42/52), was more frequently detected in the genomes from China (60/172) than in those from the USA (10/259) (**Figure 6B**).

### Geographic dissemination of major clones

To assess the phylogeographic structure of each of the dominant MRSA and MSSA high-risk clones identified in our study, we compared the genetic relatedness between isolates from the same country and from different countries. Pairwise differences in the number of SNPs from core genomes are often used to estimate the level of genetic relatedness between isolates. Comparisons of the pairwise SNP differences within MRSA-ST30 and MSSA-CC398 showed significantly different median pairwise SNP distance between genomes from the same country and genomes from different countries (*p*<0.0001, Wilcoxon test), but the interpretation adjusting for size effects shows that the magnitude of the differences is small or negligible (Cohen’s *d* value <0.30, Cliff’s delta value <0.16) **(Figure 7)**. This suggests that, despite their geographical spread, isolates from different countries are, on average, as similar to each other as are isolates from the same country, and further supports the observed frequent transmission across borders of these dominant high-risk clones **(Figures 3-6)**.

**Figure 7.**
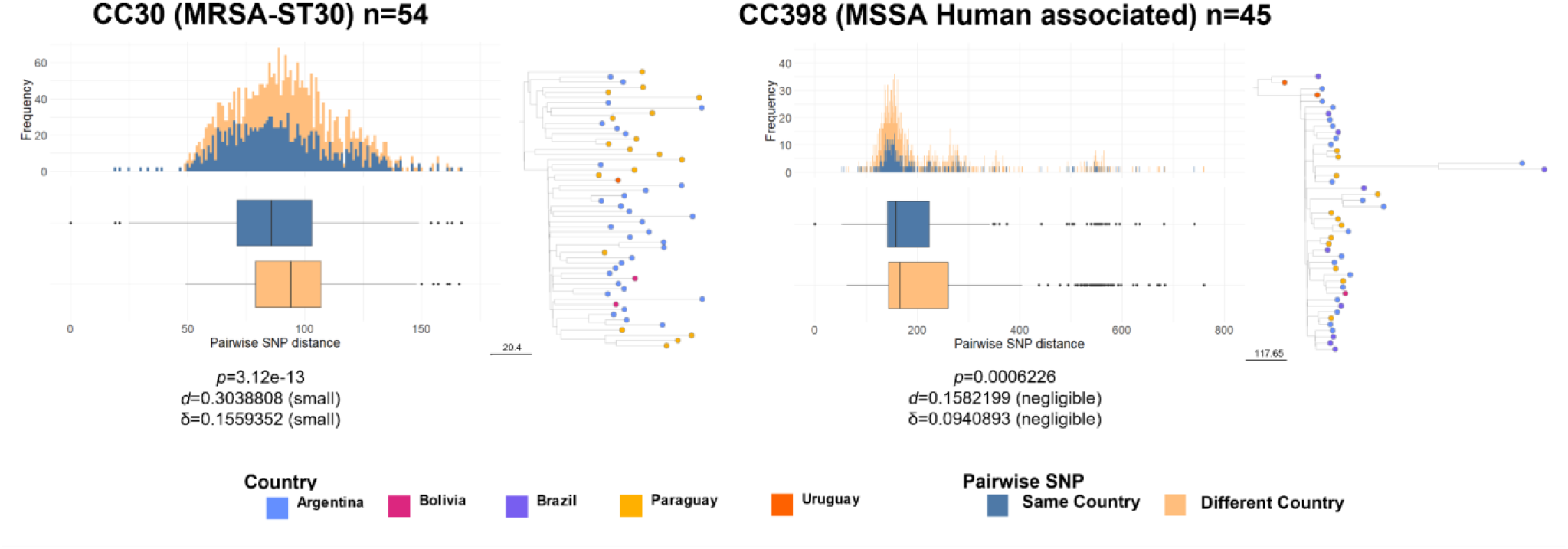
Distributions of core genome SNP differences between pairs of genomes belonging to the same clade from either the same (blue) or different (orange) countries. Pairwise SNP distance is represented as boxplots (median and the interquartile range) and a histogram of frequency for each clade. The number of genomes included is detailed in the legend for each of the main lineages analysed. Text in the bottom of each figure is an interpretation of the difference between each pair of distributions, obtained using the R package “effsize,” which applies the parametric and non-parametric effect size estimators Cohen’s *p*, and Cliff ‘s δ to the results of a Wilcoxon test (*p*). Subtrees of each main clade are represented next to each plot. Scale bar next to the tree represents the number of SNPs. Nodes (genomes included in the analysis) are coloured by country as described in the legend.

## Discussion

Our study provides a contemporary snapshot of the genetic characteristics and the epidemiology of MRSA and MSSA causing bacteremia in the southern cone of South America during 2019 through WGS and a rational sampling framework like the one used in previous surveys in Europe [16, 17]. However, the sampling framework presented several challenges. We were not always able to recruit enough participating centres to fully represent each country demographically and geographically.

The geographical representation of our survey is especially limited in Brazil, where few laboratories were enrolled, and therefore, the data unlikely reflects the entire epidemiological situation of this large country. Similarly, the coverage of the southern cone is curtailed by the absence of Chile in this first survey. In addition, almost half of the centres were unable to submit ten *S. aureus* isolates, which may represent both the constraints faced by laboratories in low-resource areas (that nonetheless participated voluntarily), and the decreasing trends in incidence of *S. aureus* bacteraemia [70]. Despite these limitations, our study is the largest genomic survey of the *S. aureus* population in South America to date, which we characterised with a resolution that enabled the identification of MRSA and MSSA lineages of public health significance in the region, informed on their geographic structure, placed them in global context, and linked them to AMR phenotypes.

Changes in *S. aureus* AMR trends have been observed in Argentina, Bolivia and Paraguay during the last 8 years [7, 71, 72] likely due to clonal replacements. Our study addresses the gaps in knowledge between phenotypic resistance and population structure by characterising the more prevalent MRSA and MSSA lineages causing bacteraemia occurring in the region during 2019. The higher diversity of MSSA lineages in South America **(Figure 1)** parallels similar findings worldwide [16, 73–77]. The most prevalent MSSA genotype (CC398-MSSA-*t1451*-*lukS/F-PV*-) carrying the *ermT* gene and *sh_fabI* in a mobile genetic element associated with triclosan resistance [69], is largely responsible for the erythromycin and clindamycin resistance rates observed in MSSA strains (29.5% and 29.1%, respectively, **Table 3, Figure 6)**, which had been rising prior our sampling period, and continued to rise post COVID-19 pandemic [71, 72]. To our knowledge, human-associated CC398-MSSA-*t1451*-*lukS/F-PV*-, is described here for the first time as an epidemic MSSA clone in our region.

Between 2006 and 2014, CC398 was only sporadically found in South America colonising and causing invasive infections in humans [78–83]. Since 2011, in Argentina and Uruguay empirical antimicrobial therapies for skin and soft tissue infections include clindamycin, trimethoprim-sulfamethoxazole or minocycline due to the high prevalence of CA-MRSA [84, 85]. On the other hand, triclosan is an antiseptic widely used in our region [86]. We hypothesise that CC398-MSSA may have recently become a prevalent pathogen in response to the selection pressure imposed by this public health measure, at least in Argentina and Uruguay, or by the indiscriminate use of triclosan, although we cannot rule out other unexplored factors. However, the prevalence of CC398 as an agent of bacteraemia prior to 2019 remains uncertain due to the paucity of surveillance studies of MSSA in South America.

Although cefazolin is a common treatment option for MSSA bacteraemia, older South American bloodstream MSSA isolates carrying BlaZ types A and C have previously exhibited a high frequency of the cefazolin inoculum effect, which is associated with treatment failure and increased mortality [83, 87]. Almost half of the MSSA genomes analysed here, including CC398, harboured BlaZ type B (113/239, 47.3%) **(Suppl Table 5)**, which does not lead this phenotype [87, 88]. However, a high proportion of genomes from other CCs carried BlaZ type A or C (90/239, 37.7%) **(Suppl Table 5)**, especially so in Bolivia (18/31, 58.1%) where CC398 is not as prevalent. Therefore, although cefazolin seems to be a viable option for the treatment of MSSA in our region, further studies are needed to understand potential country-specific differences in the cefazolin inoculum effect.

Despite *S. aureus* strains recovered from bacteremia in our study are in general not MDR **(Table 3, Figure 2B**), MRSA strains pose an additional challenge to treatment because of higher resistance rates to additional antibiotics besides beta-lactams **(Table 3)**. The MRSA sub-population was dominated by three CCs (CC30, CC5, and CC8), and in particular by three genotypes, CC30-MRSA-IVc-*t019-lukS/F-PV*+, CC5-MRSA-IV*-t002-lukS/F-PV-*, and CC8-MRSA-IVc*-t008-lukS/F-PV+*-COMER+ **(Figure 2)**.

Global phylogenetic analysis of CC30-MRSA-IVc-*t019-lukS/F-PV*+ showed a distinct epidemic of this SWP-related clone affecting the Southern Cone of South America, in particular Paraguay and Argentina, where it is relatively more prevalent than other MRSA. Our findings suggest a single introduction in South America of an MRSA-ST30 clone that disseminated throughout the continent, likely via travel routes, in agreement with previous reports [9, 47–49, 85, 89–91]. ST30-MRSA was first described in our region from Uruguay [92] and Brazil [93]. The paucity of genomes from earlier dates precludes us from establishing the entry point into the region with the current analysis.

Interestingly, genomes from this study belonging to CC30-MRSA-IVc-t019-lukS/F-PV+ were found on longer terminal branches than related genomes from 2005-2014 in the same clade, suggesting the accumulation of more genetic variability **(Figure 3)**. Our previous work [15] describes the acquisition of a premature stop codon in the *tag* gene (coding for DNA-3-methyladenine glycosylase) ancestral to this clade. The loss of function of this enzyme in charge of removal of chemically damaged DNA, might be related with the accumulation of genetic changes observed. *S. aureus* hypermutator strains, caused by defects in the methyl-mismatch repair system via mutations on the *mutSL* locus, have been previously found to play a role in AMR development during long-term persistence [94, 95]. Interestingly, the gateway of bacteraemia for 25 of 49 CC30-MRSA-IVc-t019-lukS/F-PV+ genomes are SSTI (n=22) and surgical wounds (n=3), which suggests possible long-term colonisation of the patients’ skin. In contrast, none of the other CC30-MRSA found on different clades of the CC30 tree (one CC30-MRSA-IVa-t318-lukS/F-PV+ and two CC30-MRSA-IVc-t433-lukS/F-PV+) were known to be from SSTI or surgical wounds. A possible relationship between DNA repair and the success of this MRSA lineage will be the subject of future investigation **(Figure 3)**.

The genotype CC5-MRSA-IV*-t002-lukS/F-PV-* was contextualised within the previously defined CC5-basal clade, as opposed to the CC5 Cordobés/Chilean (or CC5-I) clone once dominant in the region. The polyphyly of CC5-basal genomes was reflected on a variable repertoire of toxins [54] **(Figure 4)**. Remarkably, most of them lacked the staphylococcal superantigen-like protein *set9*/*ssl4* gene which is located within the variable region of the genomic island νSaα [96].

We placed CC8-MRSA-IVc-t008-*lukS/F-PV+* COMER+ within the epidemic USA300-SAE clone previously described for the north of South America **(Figure 5)** [10]. Importantly, we describe a variant of USA300-SAE lacking a functional capsular biosynthesis gene *capE* as the most prevalent MRSA clone in Bolivia, which likely evolved from Colombian ancestors[10]. Isolates with similar epidemiological markers were reported from human colonisation and infections in Bolivia 2010-2013, suggesting the presence of this clone in the country during the last decade [79, 97]. Our study delivers local value by demonstrating the importance of this clone in Bolivia, as well as its spread to other countries in the southern cone of South America, and link to a public genome from a travel-associated infection in Europe [97].

The lower AMR rates found in CA-MRSA strains compared to HA-MRSA **(Figure 2, Table 3)** are linked to two genotypes CC30-MRSA-IVc-t019-lukS/F-PV+ and CC8-MRSA-IVc-t008-lukS/F-PV+-COMER+ **(Figure 2)**, which are historically from a CA origin, are overrepresented in CA-MRSA compared to HA-MRSA and carry a small number of AMR determinants. Similar to reports from other regions [98–100], MDR HA-MRSA lineages previously dominant in South America (ST239-MRSA-III “Brazilian” clone, ST5-MRSA-I “Chilean/Cordobés” clone, ST105-MRSA-II “Rio de Janeiro” clone)[47, 55, 56, 101, 102] were replaced by non-MDR CA-MRSA that now seem to have fully established within the hospital environment and cause bloodstream infections. The lack of anti-restriction gene homologs previously associated with MDR [103] might partially explain (at least in part) the low levels of AMR in these strains.

In conclusion, the majority of the dominant genotypes causing bacteraemia in South America are high risk clones with a CA origin and related to global epidemic clones. Nevertheless, we demonstrate the presence of local and regional epidemics of these high risk clones within South America, e.g. of a descendant of the SWP clone (MRSA-ST30) affecting mainly Argentina and Paraguay, of a variant of USA300-SAE mainly in Bolivia, and of CC398-MSSA mainly in Argentina, Paraguay and Brazil. MRSA-ST30 and CC398-MSSA did not exhibit clear country-specific phylogenetic signals **(Figure 7)**, consistent with readily community transmission amongst colonised individuals in a region where travel between neighbouring countries is frequent.

Several of these clones lacked intact copies of key virulence genes such as *geh*/*icaD*/*tag* in MRSA-ST30, *set9* in CC5-Basal, *lukS*/*F*-PV in USA300-Early branching clades or *capE*/*selq* in USA300-SAE **(Figure 3-6)**, further supporting the added value of genomics in characterising locally circulating lineages and distinguishing them from those found in other regions/countries. This is particularly so in the light of previous work showing that absence/loss of virulence properties and the concomitant reduced toxicity could pave the way for CA clones of *S. aureus* to continue to cause invasive infections in the hospital environment [104–108]. In contrast to virulence gene loss, the sporadic acquisition of AMR determinants we observed within these CA clones could lead to the selection and expansion of new resistant clones. Taking into account previous reports from Brazil showing that CC398-MSSA-t1451 can acquire the SCC*mec* cassette [58, 80], and the widespread use of antibiotics and antiseptics during the COVID-19 pandemic, the emergence of epidemic CC398-MRSA clones with MLSb resistance is a plausible scenario.

Taken together, our findings highlight the need for regional ongoing genomic surveillance by networks such as StaphNET-SA. Structured surveys undertaken at regular time intervals could uncover the population dynamics of this pathogen through the comparison with the baseline population established in this study. We made available the assemblies from this first survey via the PathogenWatch platform to facilitate the exploration of these data and the comparison with datasets from other countries/regions. Structured surveys were conducted in Europe in the past [16, 17] and, were they to be adopted in other regions in a standardised and concerted manner, they could amount to a global picture of this pathogen that was responsible for the largest number of deaths worldwide in 2019 [109].

## Data Availability

All supporting data, code and protocols have been provided within the article or through
supplementary data files. Five supplementary figures and five supplementary tables are available
with the online version of this article.
Sequence read files for all samples used in this study have been deposited in the European
Nucleotide Archive under the project accession number PRJEB37318.

## Author contributions

S.D.G, J.V, M.S.H., and L.R. performed phenotypic characterization and data curation.

J.C. and M.S.H. performed DNA extraction.

M.K. performed management of bacterial DNA samples.

S.D.G., J.V, S.A. and M.S.H. analysed the data.

S.A. and S.D.G. designed and supervised the data analyses.

S.D.G, J.C., A.F., G.R., M.H.S., M.H., N.B.Y., A.F., M.F., R.G., T.C., G.V., D.M.A., S.A. and M.M. conceived the study.

S.D.G, J.V., S.A and M.M. wrote the original draft.

All authors reviewed, edited, and approved the final version of the manuscript.

## Acknowledgements

**StaphNET-SA consortium:** Univ. de Buenos Aires: Mollerach Marta, Vielma Jesús, Di Gregorio Sabrina, Haim María Sol, Rago Lucía.

Plataforma de Genómica y Bioinformática; ANLIS; Buenos Aires: Campos; Josefina, Poklepovich Tomás. PAHO/WHO: Marcelo Galas, Washington, USA.

Centre for Genomic Pathogen Surveillance, BDI; Univ. Oxford; United Kingdom: Argimón Silvia, Aanensen David. Hospital Privado Regional del Sur + Sanatorio del Sol: Gabriela Rubinstein; Hospital Zonal Ramón Carrillo: Sabrina De Bunder; Sanatorio San Carlos: Graciela Parsons; Hospital de Pediatría Dr. Fernando Barreyro: Martha Von Spetch/Lorena Leguizamón; Laboratorio de Alta Complejidad de Misiones (LACMI): Viviana VIllalba; Hospital SAMIC de Oberá: Cristina Gonzalez; Hospital SAMIC de El Dorado: Ana Maria Miranda; Hospital de Clínicas José de San Martín: Ángela Famiglietti; Hospital Alemán: Liliana Fernandez Cannigia; Sanatorio Guemes: Soledad Zárate; Hospital Universitario Austral: Viviana Vilches; Hospital Santojanni: Claudia Alfonso; IACA Laboratorios (Bahía Blanca): Claudio Chavez; Hospital H.I.G.A. “Dr. José Penna”: Claudio Chavez; Hospital Público Materno Infantil: Ana Berejnoi; Hospital San Bernardo: Jorgelina Mulki/Emilce Flores/Viviana Silva; Hospital Luis Lagomaggiore: Silvia Atorri; Hospital San Martín: Mariana Boleas; Clínica Universitaria Reina Fabiola: Marina Bottiglieri; Hospital de Niños Dr. Orlando Alassia: Maria Rosa Baroni/Carolina Aro; Hospital de Reconquista “Olga Stuky de Rizzi”: Carina Muchiut/Otilia Sellares; Hospital “Victor J Vilela” de Rosario: Andrea Badano/Adriana Ernst; Sanatorio Adventista del Plata: Melina Herrera/Graciela Posse; Instituto de Cardiología Juana Cabral: Laura Peña; Laboratorio de Analisis Clinicos y Bacteorologicos San Miguel: Luis Fabian Aguirre; Laboratorio Hospital Regional Rio Grande: Silvia Longoni; Hospital Delicia Concepción Masvernat: María Ofelia Moulins; Hospital Daniel Bracamonte: Marlene Castillo Cruz; Caja Nacional de Salud: Rosmery Virginia Aguilar Arispe; Hospital del Niño Manuel Ascencio Villarroel: Norah Balderrama Yarhui; Hospital General San Juan de Dios: Margoth Carola Benavidez Carrillo; Hospital Santa Bárbara: Rosario Navia; Hospital del Niño Sor Teresa Huarte Tama: Jackeline Chávez Vicker; Hospital del Niño Mario Ortiz: Blanca Machuca/Zulma García Montaño; Caja Nacional de Salud: Maria Elena Arauz; Hospital Materno Infantil Boliviano Japonés: Miriam Velez; Hospital in Copacabana Area: Agnes Figueiredo; Hospital in Meier Area: Agnes Figueiredo; Hospital Regional Joinville: Alessandro Silveira; Hospital Universitário de la Universidad de São Paulo: Nilton Lincopan; Hospital Universitário Polydoro Ernani de São Thiago - UFSC: Tahis Sincero; Natal Garrison Hospital, Brazilian Army: Nilton Lincopan; Hospital Regional de Ciudad del Este: Nancy Segovia Coronel; Hospital Central de IPS: Gladys Velázquez; Hospital General Pediátrico: Noemí Zárate; Hospital Nacional de Itaguá: Gloria Gomez; Instituto de Medicina Tropical (IMT): Matilde Outeda; Instituto Nacional de Enfermedades Respiratorias (INERAM): Rossana Franco; Instituto de Previsión Social Periférica (IPS-NANAWA): Nancy Melgarejo; Laboratorio privado Diaz Gill: Sofia Busignani; Hospital General de Luque: Rosana Ortiz; CAMEC: Noelia Burger; COMERO: Rosina Servetto; CAMEDUR: Claudia Gutierrez; cHPR: Gabriela Algorta; Hospital Maciel: Antonio Galiana; CASMU: Eugenia Torres; Sociedad Médico-Quirúrgica: Paula Arralde; Hospital de Treinta y Tres: Luis Jorge

