## Supplementary Figures for "Genomic epidemiology of *S. aureus* isolated from bloodstream infections in South America during 2019 supports regional surveillance"

**Suppl Fig. 1.** Sampling map of StaphNET-SA study surveillance sites. Coloured pie charts depict the distribution of MRSA (red) and MSSA (light blue); pie size is scaled according to the number of isolates collected at each site.

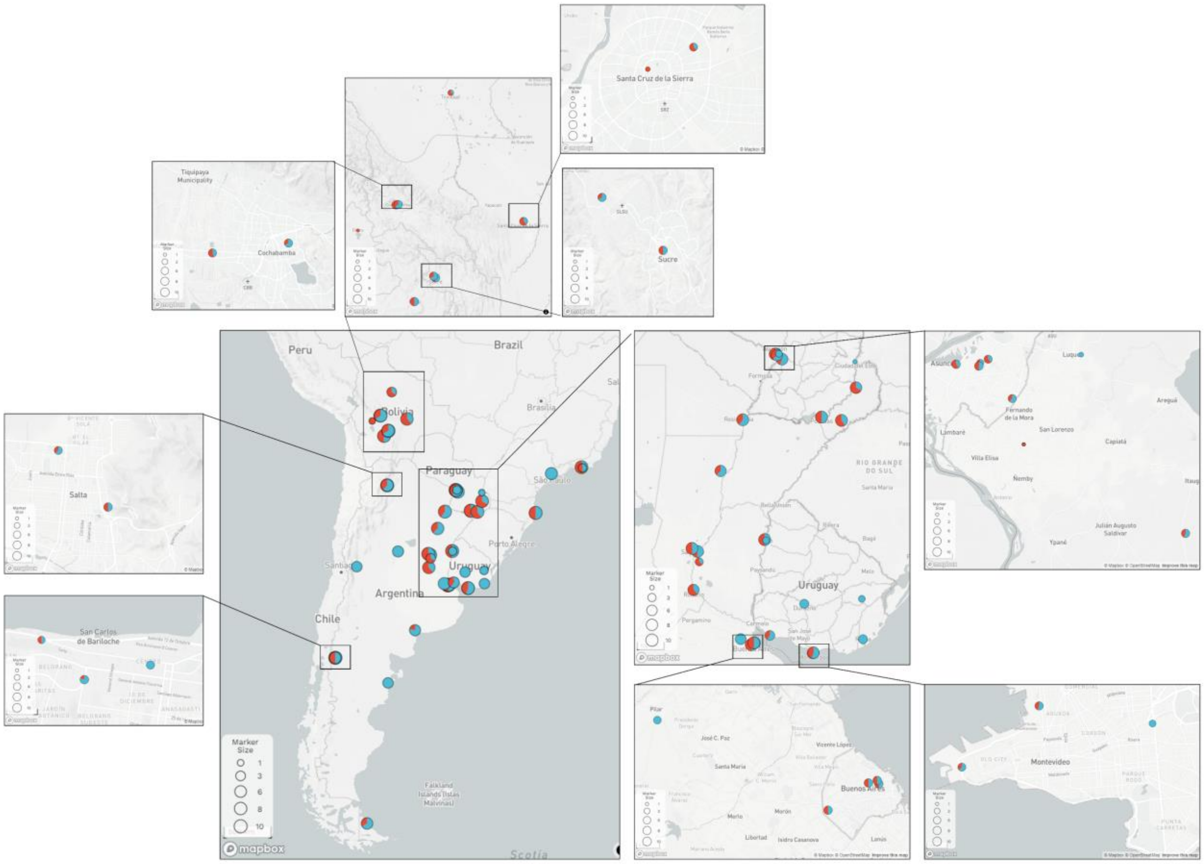

**Supplementary Fig. 2**. Diagram flow showing confirmation and quality control performed on *S. aureus* isolates and genomes in this study.

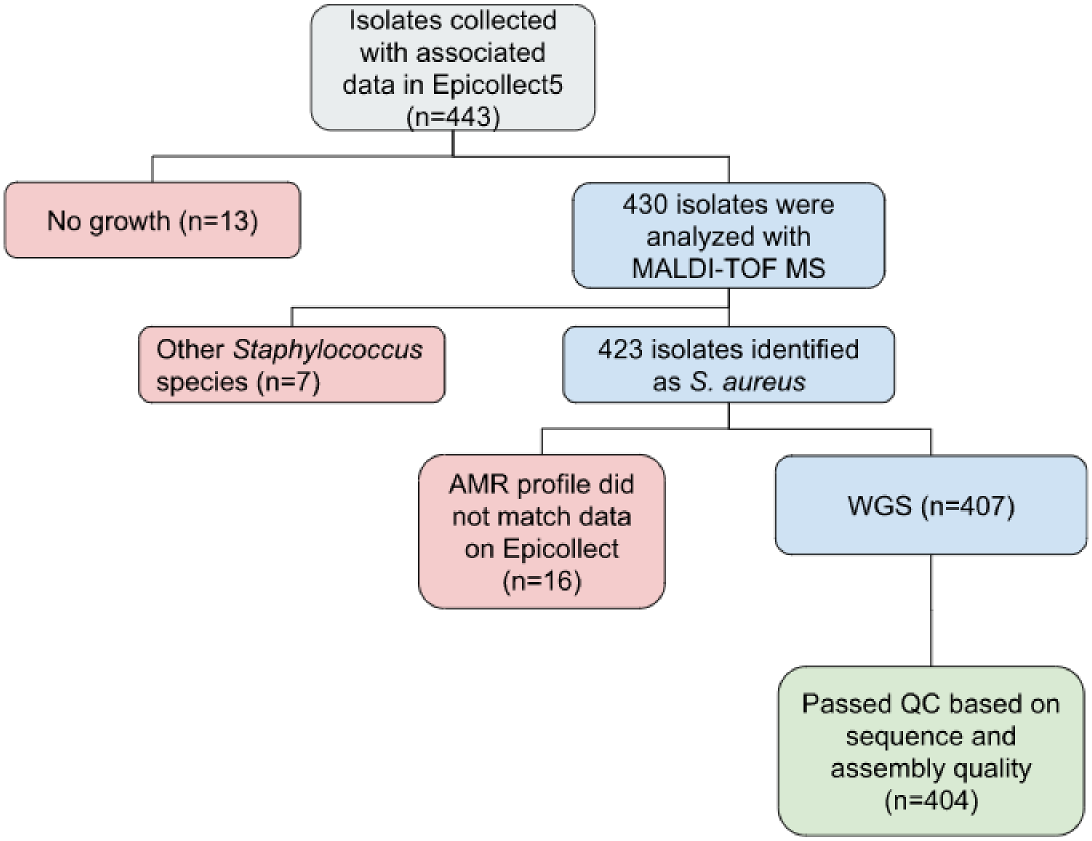

**Supplementary Fig. 3**. Distribution of clonal complexes in the 404 *S. aureus* genomes by MRSA or MSSA. STs comprising less than 3 genomes are grouped under “Others”. Bars are coloured as described in the legend.

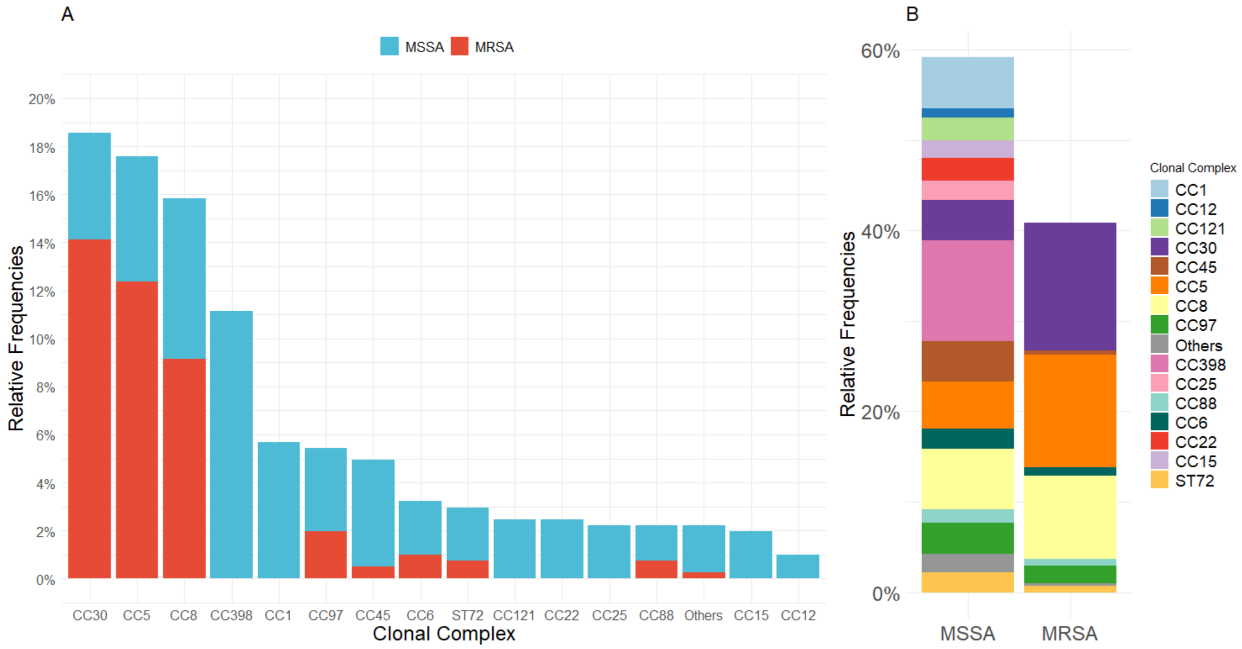

**Suppl Fig.4.** AMR determinants and AMR phenotypes. Maximum Likelihood tree of 404 genomes inferred from 156868 SNP sites identified on 2182 core genes (Panaroo) with RAxML. Midpoint rooted. 500 bootstrap replicates. Tree nodes and blocks are coloured as described in the legend.

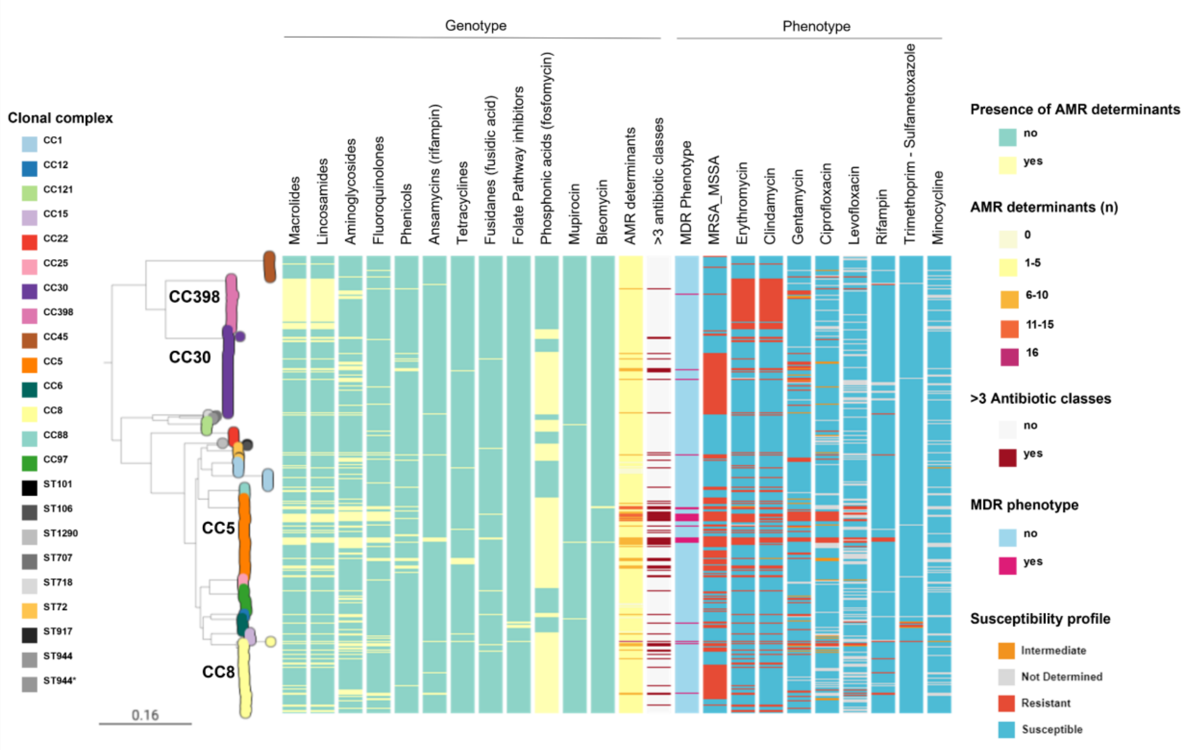

**Suppl Fig. 5.** Geographic distribution of USA300 Early Branching clades. Leaf nodes are coloured by Country. Coloured blocks represent the presence of an intact genetic determinant: virulence gene (violet). Country and SCC*mec* type colours are described in the legend. For both trees, the outgroup is omitted, and scale bars represent the number of single nucleotide polymorphisms (SNPs) per variable site.

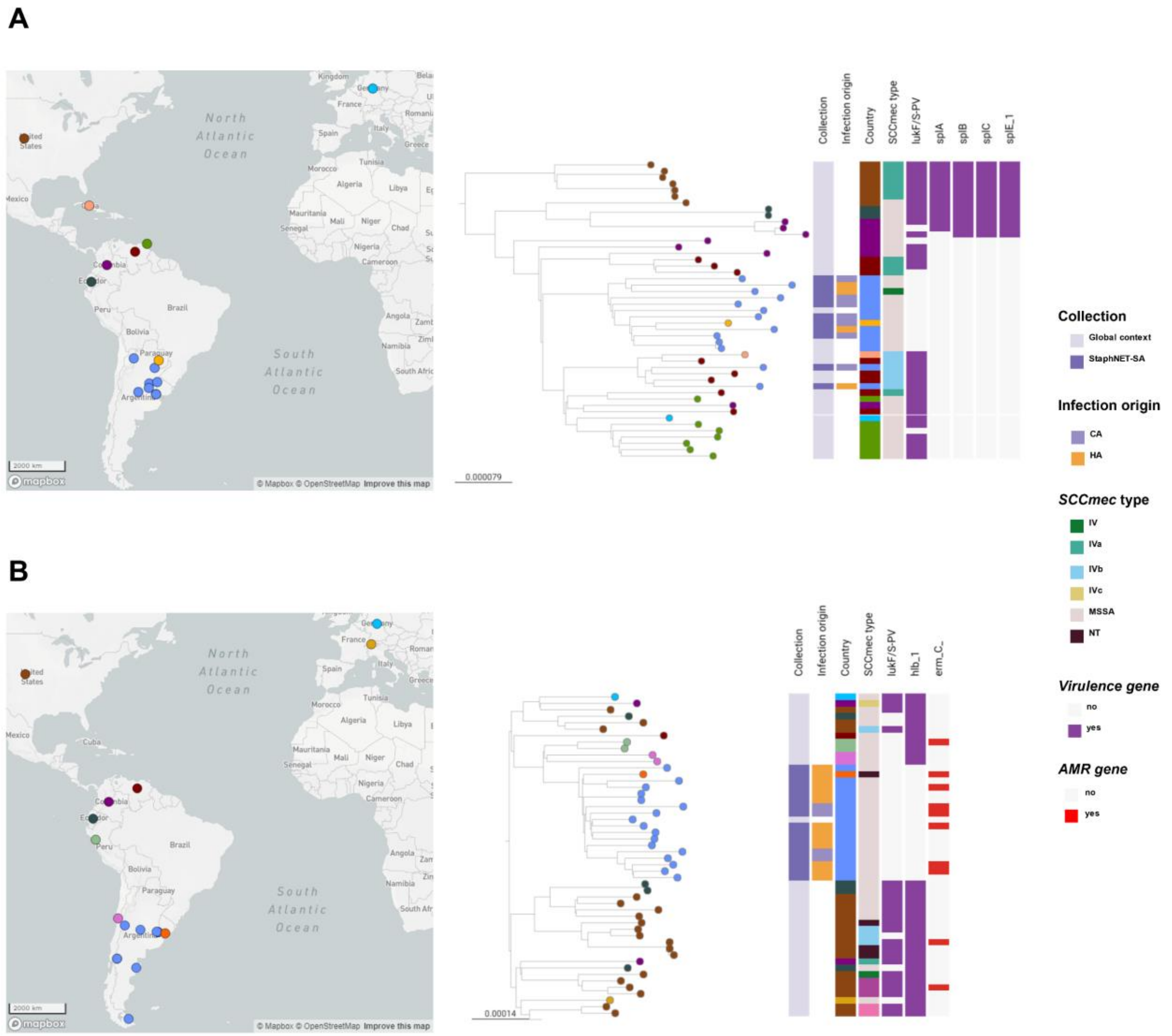
